# Tat_BioV: Tattoo ink exposure and biokinetics of selected tracers in a short-term clinical study of 24 subjects

**DOI:** 10.1101/2024.08.16.24311916

**Authors:** Susanne Kochs, Sandra Schiewe, Milena Foerster, Kathrin Hillmann, Claudia Blankenstein, Martina C. Meinke, Josephine Kugler, David Kocovic, Andreas Luch, Ulrike Blume-Peytavi, Ines Schreiver

**Affiliations:** German Federal Institute for Risk Assessment (BfR), Department of Chemical and Product Safety, Berlin, Germany; International Agency for Research on Cancer (IARC), Environment and Lifestyle Epidemiology Branch, Lyon, France; Charité-Universitätsmedizin Berlin, Corporate Member of Freie Universität Berlin and Humboldt Universität zu Berlin, Department of Dermatology, Venereology and Allergology, Clinical Research Center for Hair and Skin Science, Berlin, Germany; Charité-Universitätsmedizin Berlin, Corporate Member of Freie Universität Berlin and Humboldt Universität zu Berlin, Department of Dermatology, Venereology and Allergology, Center of Experimental and Applied Cutaneous Physiology, Berlin, Germany; Institute for Medicines and Medical Devices of Montenegro, Center for Inspection Supervision and Market Control, Podgorica, Montenegro

## Abstract

**Background:** About one-fifth of people in industrialised countries are tattooed, potentially putting them at risk of exposure to possible carcinogenic or otherwise harmful substances. Health risks are directly correlated with the amounts of substances introduced, yet reliable data on the systemic exposure to tattoo inks are lacking.

**Objectives:** This study aims to determine the exposure to soluble tattoo ink ingredients and their excretion within 24 hours after tattooing. Comparative *in vivo* and *in vitro* experiments were conducted to determine the change in metabolite exposure between tattooing and oral exposure.

**Methods:** In a clinical study, 24 subjects were tattooed with black or red tattoo ink to which the tracer substances potassium iodide, 4-aminobenzoic acid (PABA) and 2-phenoxyethanol (PEtOH) had been added to mimic known hazardous substances found in tattoo inks. Tracers and their metabolites were quantified in blood, urine, ink, and consumables pre- and post-tattooing. Tattooed skin area was determined using picture analysis. PABA metabolism upon tattooing was compared to peroral administration. Skin fibroblasts and macrophages were tested *in vitro* for their ability to metabolise PABA.

**Results:** All tracers or their metabolites were identified in urine; iodide and the PABA metabolite 4-acetamidobenzoic acid (ACD) were identified in plasma. The worst-case scenario for systemic ink exposure was estimated to be 0.31 g of ink per tattoo session (75^th^ percentile). Peroral administration resulted in lower levels of ACD than tattooing. Fibroblasts and macrophages were capable of converting PABA into ACD.

**Discussion:** Our results are the first human *in vivo* data on soluble tattoo ink ingredients and suggest that the overall exposure might be lower than the estimates previously used for regulatory purposes. In addition, the first-pass effect by skin metabolism leads to an altered metabolite profile compared to oral exposure. Skin metabolism might also contribute to detoxification of certain carcinogenic substances through *N*-acetylation.

## Introduction

The high prevalence of body tattooing and its facial equivalent, the latter better known as permanent make-up, has already raised health concerns for decades.^1^ Between 17 and 31.5% of all people across Europe and the United States are tattooed.^2,3^ The European REACH (Registration, Evaluation, Authorisation and Restriction of Chemicals) restriction on tattoo and permanent make-up inks bans more than 4000 substances due to their hazard potential, thus making it the strictest regulation worldwide.^4^ Substances with toxic potential found in tattoo inks include heavy metals (e.g., chromium, nickel, cobalt), primary aromatic amines (PAAs) and polycyclic aromatic hydrocarbons (PAHs), which may have allergic and/or carcinogenic potential.^5–12^ While acute adverse reactions occur promptly after tattooing and are therefore relatively easy to correlate with them, long-term effects of tattoos, such as cancer and immunotoxicity, are not easily linked.^13^ Exposure quantification is crucial to calculate safe limits and to assess population-wide risks associated with intentionally or non-intentionally added hazardous substances in tattoo inks.

During tattooing, insoluble pigments and soluble co-formulants are introduced into the dermal layer of the skin. Previous studies as well as clinical observations have shown that pigments can be transported to regional lymph nodes of tattooed individuals,^14^ indicating the possibility of further migration towards more distant organs. Since pigments remain in the body, long-term exposure has to be considered, whereas water soluble ingredients are likely to be metabolised and excreted rapidly. In this case, a short-term, acute exposure period can be assumed. Exposure through tattooing is determined by the amount of ink applied per surface area of the tattooed skin. Several studies were conducted to determine exposure levels, including weighing *ex vivo* skin before and after tattooing^15^ and quantifying pigments after tattooing of *ex vivo* skin.^16^ However, these data primarily refer to the initial pigment deposition rather than the soluble fraction administered, and they vary greatly (0.6 – 9.5 mg_pigment_/cm^2^).^16^ In 2017, the European Chemicals Agency (ECHA) estimated an exposure level of 14.36 mg_ink_/cm^2^ based on a study by Engel and colleagues.^16^ For a tattooed area of 300 cm^2^, this would result in an equivalent to 4.308 g of ink per session.^17^ However, these were *ex vivo* generated data and therefore do not consider the actual absorption of the soluble ink fraction into the body. Thus, their relevance for real-life exposure estimation remains elusive.

As tattooing is a unique kind of exposure route, comparison of metabolic profiles with peroral dosing – an administration route often used in toxicity studies – is of high interest. Skin metabolism was previously described as detoxifying for carcinogens such as PAAs.^18^ A different metabolic profile might therefore lead to increased or reduced adverse effects despite a similar systemic dose. Main driver of this detoxification is the *N*-acetylation of PAAs by keratinocytes.^18^ However, gene expression of *N*-acetyltransferase 1 (NAT1) was also found in fibroblasts.^19,20^ Fibroblasts are the most common cell type in the dermis and tattoo pigments reside predominantly in fibroblasts and macrophages.^21,22^

In the present study, we used an *in vivo* human quasi-experimental design to derive a reasonable worst-case exposure scenario that may be applied in future risk assessments. As most of the potentially harmful tattoo ink ingredients cannot be used in human experimental exposure studies due to their hazard potential, we added tracer substances (potassium iodide, 4-aminobenzoic acid (PABA), and 2-phenoxyethanol (PEtOH)) to tattoo inks, as hazard-free alternatives. To estimate the short-term exposure and biodistribution of tattoo inks, the tracers were quantified in blood, urine and consumable products in contact with ink of 24 subjects by using previously validated analytical methods.^23^ The tattooed body surface per session was derived by digital picture analysis. In addition, skin and peroral metabolism of PABA were investigated *in vitro* and *in vivo*, respectively.

## Methods

### Study design

The single-arm study was conducted at the Clinical Research Center for Hair and Skin Science, Charité-Universitätsmedizin Berlin (Berlin, Germany) from November 2021 to September 2022. A total of 24 subjects were tattooed by professional tattoo artists with different tattoo inks (14 black, 10 red) spiked with the tracers iodide, PABA and PEtOH. These tracers were selected based on their physico-chemical similarities to tattoo-associated substances, whilst having a low toxicological profile and also being used in drug products (details cf. *Supplemental Material 1*).^23–30^ Iodide was chosen for its straightforward quantifiability for the determination of applied ink, PABA has structural similarities to PAAs, and PEtOH was selected as a potential preservative in tattoo inks that has similarities to the more commonly used benzyl alcohol. Tracers were added to commercial tattoo inks at a clinical pharmacy unit under sterile conditions (cf. *Supplemental Material 1*). Subjects and tattoo artists were recruited through flyers and social media. The inclusion criteria for subjects were: healthy, male, 18 – 45 years of age, 60 – 100 kg, autonomous wish for a tattoo (tattooed area ± 30%: black ∼ 300 cm^2^, red ∼ 100 cm^2^), and at least one already existing tattoo. The design of the tattoo and the tattoo artist were of subject’s choice. Exclusion criteria included health restrictions, intake of certain drugs or dietary supplements containing the tracers or that might otherwise affect the study, and unwillingness to follow the behavioural and dietary rules (*Supplemental Material 2*). Since PEtOH can be present in cosmetic products, subjects were instructed to use specified cosmetic products provided for the study period only.

Participation in the study involved 3–4 appointments for each subject. Preliminary examinations included parameters such as age, height, weight, body fat (three-site measurement technique according to Jackson and Pollock^31^) and blood parameters to determine liver, kidney and thyroid function. Subjects were instructed on usage of urine canisters, dietary rules and provided with cosmetic products.

Urine was collected autonomously by subjects for 48 hours with container changes at specific time intervals (*Figure 1*). Blood samples were collected at specific time points (*Figure 1*) at the study centre using 5 ml citrate-phosphate-dextrose-adenine (CPDA) blood collection tubes. During and at the end of the tattooing process, all consumables and items that had come into contact with the ink were collected in designated bags. The samples were then transported to the German Federal Institute for Risk Assessment (Bundesinstitut für Risikobewertung, BfR, Berlin, Germany) and analysed.

**Figure 1.**
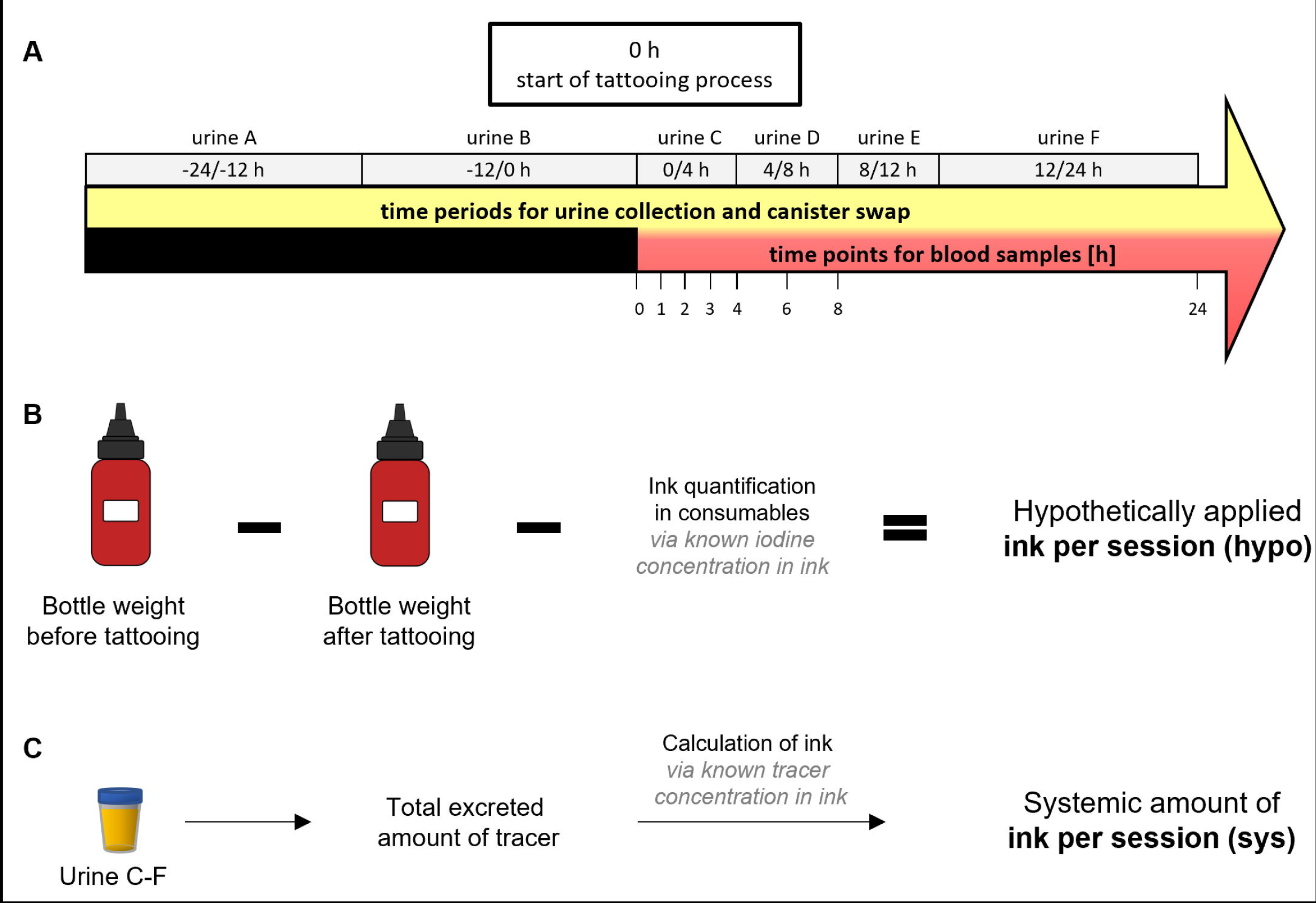
Study sampling and exposure calculation. (A) Overview of sampling times for blood and urine during the 48 h–study period. (B) Calculation of the hypothetical ink amount used per session (hypo) based on ink weighing and iodine quantification in the consumables. (C) Calculation of the systemic ink amount per session (sys) calculated by excreted tracers in urine C-F. Background levels in urine A–B were subtracted.

### Ethics approval

The study was approved by the ethics committee of the Charité – Universitätsmedizin Berlin (Berlin, Germany) under the proposal number EA4/085/21. Informed consent was obtained from all subjects before study participation. Written consent from subjects and artists was obtained for the photographs of tattoos shown. Artist copyright was indicated as requested. The study was registered in the German Clinical Trials Register under DRKS00026022 including a detailed study protocol.

### Analysis of plasma, urine, tattoo inks and consumables

Blood samples were stored and transported at 2–8 °C before processing to plasma via centrifugation at 2500 rcf for 15 min (centrifuge 5804 R, Eppendorf, Hamburg, Germany). Due to canister size, urine samples were kept at room temperature by subjects and stored and transported at 2–8 °C upon arrival at the study centre. Urine volume was determined upon arrival at the BfR.

Quantification methods of all tracers were validated according to a guideline of the European Medicines Agency with additional parameters according to an ICH guideline.^23,32,33^

For iodine quantification, plasma and urine samples as well as ink samples and consumables were analysed using inductively coupled plasma mass spectrometry (ICP-MS) after sample preparation as previously described.^23^ The method is described in detail in *Supplemental Material 3*. Consumables were extracted with 1 l ultrapure water for 2 x 30 min on every side to extract iodide while shaking (250 rpm) before addition of the internal standard.

PABA, PEtOH and their metabolites were analysed in plasma, urine and ink using high performance liquid chromatography coupled to a quadrupole time-of-flight mass spectrometer (HPLC-QTOF-MS) as previously described.^23^ Isotope-labelled internal standards were utilised for quantification against a calibration curve in the corresponding matrix (plasma or urine). In brief, sample preparation included addition of internal standards, protein precipitation by ice-cold acetonitrile (Carl Roth GmbH & Co, Karlsruhe, Germany, catalogue number: HN40.2) and centrifugation and further dilution with ultrapure water. In addition, alkaline hydrolysis was performed to determine the total PABA content in urine. For the analysis of 2-phenoxyacetic acid (PAc) in urine samples a standard addition method was used. The method is described in more detail in *Supplemental Material 3*. The cosmetic products used by the subjects were screened to confirm absence of PEtOH. No relevant concentrations were detected (*Supplemental Material 4*).

### Picture analysis

Digital photographs of the tattooed area were taken from a straight angle under appropriate light conditions. A measurement device was placed next to the tattoo for calibration purposes. Total tattooed body surface in cm^2^ was derived by digital picture analysis using the open-source software FiJi/ImageJ (version 2.3.0/1.53q) and its plugin Trainable Weka Segmentation. This method has already been previously applied for analysis of tattooed body surface.^34^

### Determination of tattoo ink, urine recovery and exposure

The amount of ink applied to the skin was calculated using two approaches (*Figure 1B,C*). Firstly, hypothetical use of ink per session (hypo) was derived by iodine quantification from consumables. The tattoo ink bottles were weighed before and after tattooing and the calculated amount of ink in the consumables was subtracted from the mass difference to obtain the amount of ink applied to the skin. Secondly, systemic exposure to ink components per session (sys) was calculated from the excreted amount of PABA or iodine in urine C-F. Since iodine has a physiological background and some individuals have a minor 4-acetamidobenzoic acid (ACD) background depending on their diet, urine A–B collected in the 24 hours before tattooing was subtracted as subject specific background. Both, hypo and sys, were divided by the derived tattooed area from the picture analysis to obtain the amount of ink per cm^2^.

For the urine recovery, the amount of each tracer excreted within 24 hours (urine C–F) was divided by ink per session (hypo). Additionally, the mean recovery of all tracers was determined for every subject.

### Data analysis

Graphs were created using GraphPad Prism Version 10.1.2 (GraphPad Software, Boston, MA, USA) and R (version 4.3.1). The standard deviation used in Microsoft Excel LTSC MSO (Redmond, WA, USA) was STDEV.P, as we aimed to display the deviation within our study population.

### PABA metabolism after oral uptake

To compare peroral and intradermal administration of substances by tattooing, three healthy volunteers took 50 mg of PABA supplement. Urine was collected according to the tattoo study (*Figure 1A*) and analysed using the same HPLC-QTOF-MS method.

### PABA metabolism in cell culture

As PAAs and PABA can be metabolised by skin, particularly through *N-*acetylation,^35,36^ the metabolism of PABA was investigated in pooled human dermal fibroblasts (HDFp) from CELLnTEC advanced cell systems AG (Bern, Switzerland, catalogue number: HDfp, lot: MC1904099) and monocyte-derived macrophages (MDM) isolated from buffy coats of three different donors as previously described.^37^ The cultivation of cells is detailed in *Supplemental Material 5*. For the experiments, cells were seeded in 12-well plates (HDFp: 8 × 10^4^ cells/well and MDM: 8.75 × 10^5^ cells/well) and incubated for 24 hours before treatment with different concentrations of PABA (0, 0.1, 1, and 10 µg/ml) dissolved in Dulbecco’s Modified Eagle’s Medium/F12 1:1 (DMEM/F12) with L-glutamine, 1.2 g/l NaHCO_3_ but without phenol red from PAN-Biotech GmbH (Aidenbach, Germany, catalogue number: P04-41650). After another 24 hours of incubation, the cell culture medium was collected and analysed using the above mentioned HPLC-QTOF-MS method.

## Results

### Quality control of study inks

All inks used in the study were analysed to confirm the absence of PEtOH. The concentrations of the tracers after their addition were determined and were all within a limit of ± 15% of the nominal tracer concentration. All analytical data, corrections (e.g., concentration of iodine in the ink had to be corrected for one subject and the concentrations of all tracers for one other subject), and justifications are reported in the supplements (*Supplemental Excel File*).

### Study subjects and deviations

The age of the subjects ranged from 22 to 43 years (median 32.5 years) and the body weight from 62 to 98 kg (median 80.5 kg). Height and body fat were 161 – 193 cm (median 181.5 cm) and 7.73 – 36.58% (median 18.38%), respectively. No dermatological observations other than normal skin reaction after tattooing were reported. Minor laboratory deviations were noted but judged by the study physician as clinically non-relevant. Other deviations included coffee consumption, intake of ibuprofen the day before tattooing, difficulties during blood sampling, deviations from the sampling times and the ink bottle falling over during tattooing. The spilled ink was cleaned up with wipes and sent in a separate bag which was analysed together with other consumables with ink residues. All subject data and deviations are reported in detail in the supplements (*Supplemental Excel File*).

### Ink exposure and urine recovery of tracers

Data sets of two subjects (Black 9, Red 4) could not be fully included due to unsuccessful quantification of iodine in the consumables. They could therefore not be used to quantify the hypothetically applied ink per session (hypo). The PEtOH recovery for subject Red 8 was 425.75% and therefore significantly higher than the applied PEtOH concentration. It is possible that cosmetic products containing PEtOH were used during the study instead of the cosmetic products provided. However, the use of other cosmetic products was not reported. This value was therefore excluded from the calculations. It was also necessary to exclude PEtOH data of subject Black 12, since exclusion of several calibration points was necessary, leading to unreliable quantification (*Table 1*).

**Table 1.**
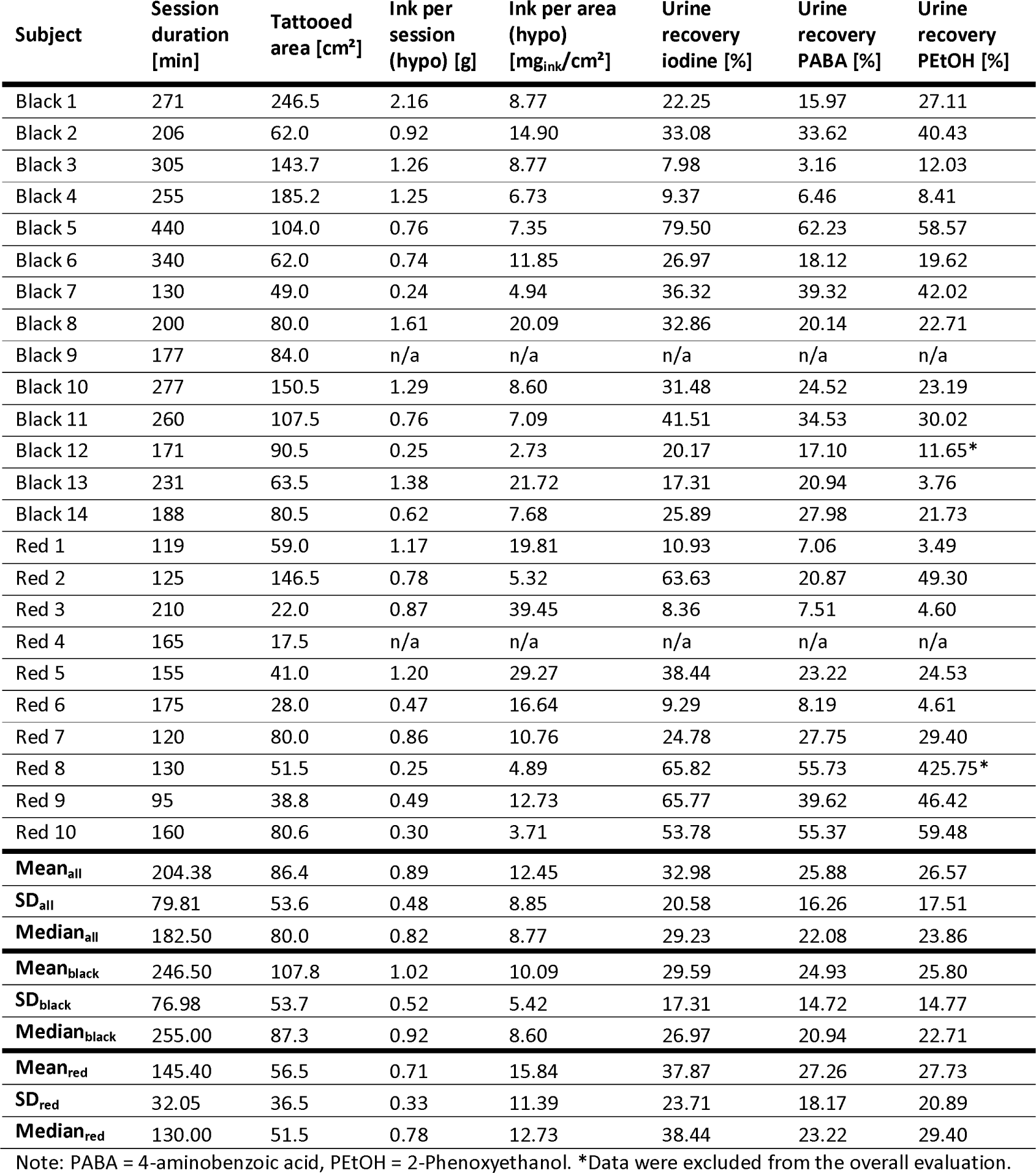
Summary of study results. Total ink and related values are based on hypothetically (hypo) applied ink dependent on iodine quantification in consumables and ink weight.

| Subject | Session duration [min] | Tattooed area [cm<sup>2</sup>] | Ink per session (hypo) [g] | Ink per area (hypo) [mg<sub>ink</sub>/cm<sup>2</sup>] | Urine recovery iodine [%] | Urine recovery PABA [%] | Urine recovery PEtOH [%] |
| --- | --- | --- | --- | --- | --- | --- | --- |
| Black 1 | 271 | 246.5 | 2.16 | 8.77 | 22.25 | 15.97 | 27.11 |
| Black 2 | 206 | 62.0 | 0.92 | 14.90 | 33.08 | 33.62 | 40.43 |
| Black 3 | 305 | 143.7 | 1.26 | 8.77 | 7.98 | 3.16 | 12.03 |
| Black 4 | 255 | 185.2 | 1.25 | 6.73 | 9.37 | 6.46 | 8.41 |
| Black 5 | 440 | 104.0 | 0.76 | 7.35 | 79.50 | 62.23 | 58.57 |
| Black 6 | 340 | 62.0 | 0.74 | 11.85 | 26.97 | 18.12 | 19.62 |
| Black 7 | 130 | 49.0 | 0.24 | 4.94 | 36.32 | 39.32 | 42.02 |
| Black 8 | 200 | 80.0 | 1.61 | 20.09 | 32.86 | 20.14 | 22.71 |
| Black 9 | 177 | 84.0 | n/a | n/a | n/a | n/a | n/a |
| Black 10 | 277 | 150.5 | 1.29 | 8.60 | 31.48 | 24.52 | 23.19 |
| Black 11 | 260 | 107.5 | 0.76 | 7.09 | 41.51 | 34.53 | 30.02 |
| Black 12 | 171 | 90.5 | 0.25 | 2.73 | 20.17 | 17.10 | 11.65* |
| Black 13 | 231 | 63.5 | 1.38 | 21.72 | 17.31 | 20.94 | 3.76 |
| Black 14 | 188 | 80.5 | 0.62 | 7.68 | 25.89 | 27.98 | 21.73 |
| Red 1 | 119 | 59.0 | 1.17 | 19.81 | 10.93 | 7.06 | 3.49 |
| Red 2 | 125 | 146.5 | 0.78 | 5.32 | 63.63 | 20.87 | 49.30 |
| Red 3 | 210 | 22.0 | 0.87 | 39.45 | 8.36 | 7.51 | 4.60 |
| Red 4 | 165 | 17.5 | n/a | n/a | n/a | n/a | n/a |
| Red 5 | 155 | 41.0 | 1.20 | 29.27 | 38.44 | 23.22 | 24.53 |
| Red 6 | 175 | 28.0 | 0.47 | 16.64 | 9.29 | 8.19 | 4.61 |
| Red 7 | 120 | 80.0 | 0.86 | 10.76 | 24.78 | 27.75 | 29.40 |
| Red 8 | 130 | 51.5 | 0.25 | 4.89 | 65.82 | 55.73 | 425.75* |
| Red 9 | 95 | 38.8 | 0.49 | 12.73 | 65.77 | 39.62 | 46.42 |
| Red 10 | 160 | 80.6 | 0.30 | 3.71 | 53.78 | 55.37 | 59.48 |
| **Mean<sub>all</sub>** | 204.38 | 86.4 | 0.89 | 12.45 | 32.98 | 25.88 | 26.57 |
| **SD<sub>all</sub>** | 79.81 | 53.6 | 0.48 | 8.85 | 20.58 | 16.26 | 17.51 |
| **Median<sub>all</sub>** | 182.50 | 80.0 | 0.82 | 8.77 | 29.23 | 22.08 | 23.86 |
| **Mean<sub>black</sub>** | 246.50 | 107.8 | 1.02 | 10.09 | 29.59 | 24.93 | 25.80 |
| **SD<sub>black</sub>** | 76.98 | 53.7 | 0.52 | 5.42 | 17.31 | 14.72 | 14.77 |
| **Median<sub>black</sub>** | 255.00 | 87.3 | 0.92 | 8.60 | 26.97 | 20.94 | 22.71 |
| **Mean<sub>red</sub>** | 145.40 | 56.5 | 0.71 | 15.84 | 37.87 | 27.26 | 27.73 |
| **SD<sub>red</sub>** | 32.05 | 36.5 | 0.33 | 11.39 | 23.71 | 18.17 | 20.89 |
| **Median<sub>red</sub>** | 130.00 | 51.5 | 0.78 | 12.73 | 38.44 | 23.22 | 29.40 |

Note: PABA = 4-aminobenzoic acid, PEtOH = 2-Phenoxyethanol. \*Data were excluded from the overall evaluation.

The average ink quantity per session (hypo) was 0.89 ± 0.48 g for both colours combined, 1.02 ± 0.52 g for black and 0.71 ± 0.33 g for red (*Table 1, Figure 2A*). The tattooed areas were subjectively estimated to be around 300 cm^2^ for black and 100–300 cm^2^ for red, whereby non-tattooed, empty spaces were included in the area. This was a prerequisite in subject recruitment, since we aimed for larger tattoos to reach detectable amounts of tracers in blood and urine. However, as demonstrated by Foerster et al.,^34^ the actual tattooed areas are often smaller than the estimated areas. We therefore used the same picture analysis method to calculate the area of the tattooed surface. The analysis of the tattooed area per session resulted in an average size of 86.4 ± 53.6 cm^2^ for both colours, with the average being 107.8 ± 53.7 cm^2^ for black tattoos and 56.5 ± 36.5 cm^2^ for red tattoos (*Table 1, Figure 2B*). The estimated exposure to tattoo ink per skin area (hypo) was therefore 12.45 ± 8.85 mg_ink_/cm^2^, with 10.09 ± 5.42 mg_ink_/cm^2^ for black and 15.84 ± 11.39 mg_ink_/cm^2^ for red (*Table 1, Figure 2C*).

**Figure 2.**
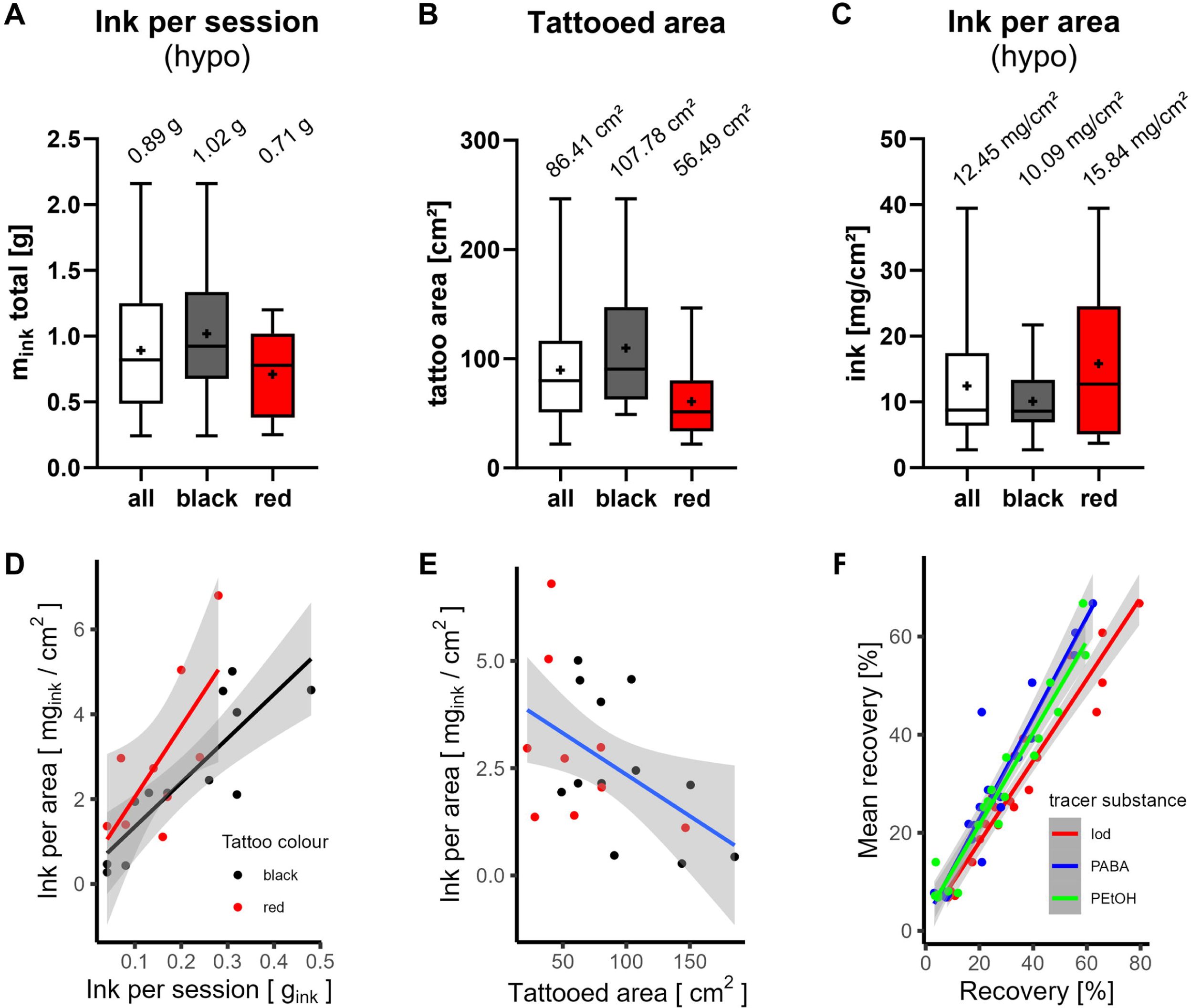
(A–C) Comparison of ink per session (hypo), tattooed skin area and ink per surface of all subjects (n = 22), ink per session calculated from mass difference of ink bottle weight and subtracted ink residues from consumables. The box and whisker plots show median (line) and mean (+) for all, black or red tattooed subjects. (D) The amount of tattoo ink per area is positively correlated with the ink use per session (systemic, sys, based on excretion of 4-aminobenzoic acid, PABA) and is higher for red tattoos. (E) Ink per area (sys, PABA) is negatively correlated with the tattooed area. (F) Correlation plot of tracer recovery to mean recovery calculated for each subject.

Black ink was used for contouring, shading and for monochromatic tattoos, whereas red ink was often used to fill larger continuous areas. In some cases, outlines or other shades were done before start of tattooing with spiked ink. The ink used for this purpose did not contain tracers and the start of blood sampling was aligned with the start of tattooing with spiked ink. Tattoo aftercare varied between tattoo artists. In one case a wound dressing was applied to a red study subject and displayed exudation of black and red ink (*Figure 3*).

**Figure 3.**
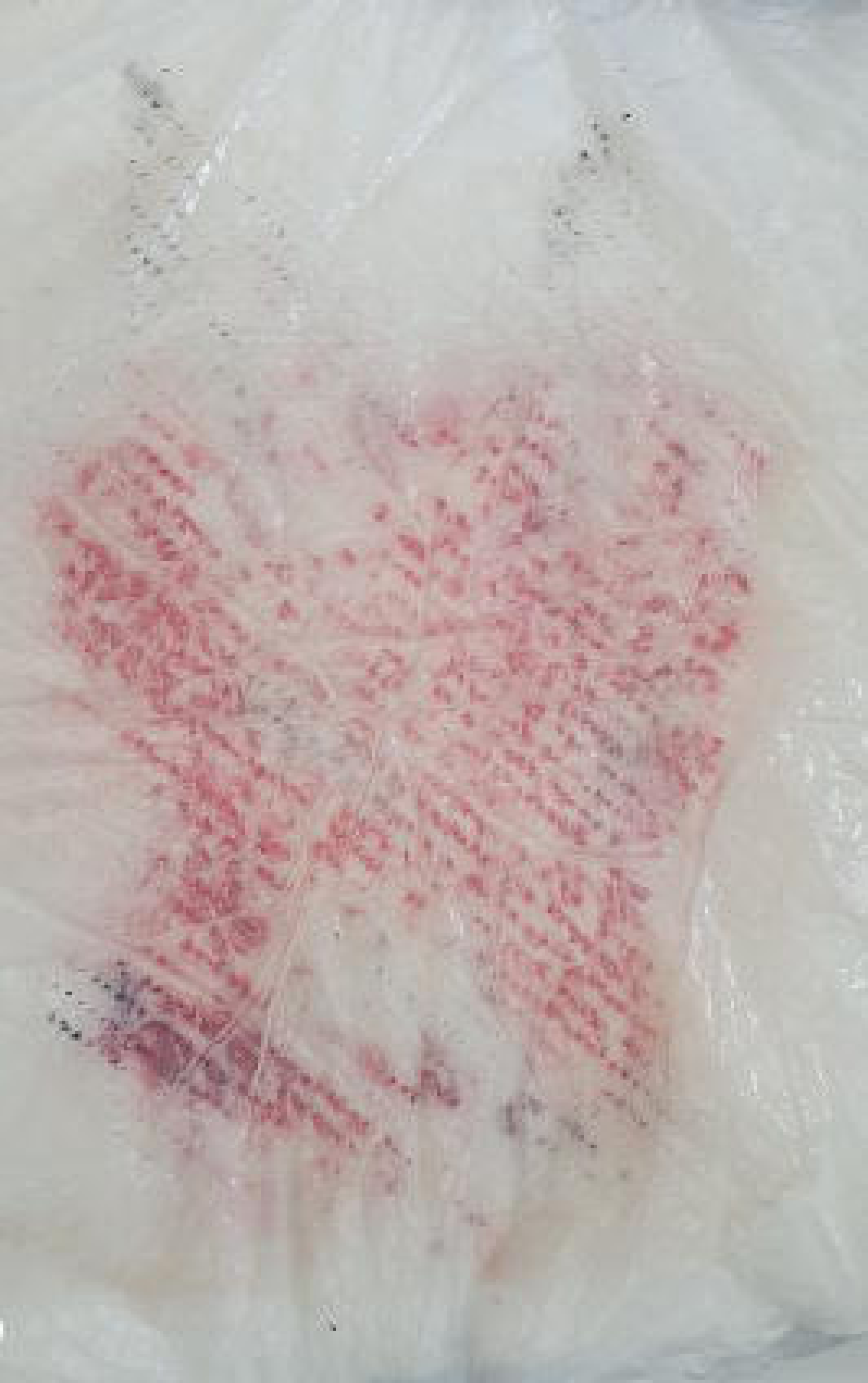
Pictures of tattoos from the study. Wound dressing of a red study subject displays exudation of black and red tattoo ink within the first hours after tattooing.

### Adjustments for absorbed fraction of tracers

The recovery of all three tracers after 24 hours was calculated by dividing the sum of excretion of each tracer in urine C-F by the initial amount of tracer in the ink per session (hypo). Although the average recoveries for all three tracers within each individual study subject were similar, the tracer recoveries for all 24 subjects were unexpectedly low and varied between 3.16 – 79.50% (*Table 1, Figure 2F*). Therefore, we re-evaluated the data with different approaches.

Firstly, the data set was reduced by excluding subjects with a mean urine recovery of < 25% (n = 13), since extremely low recoveries are most likely error prone (*Figure S1A–C*). The average amount of ink applied was 0.78 g for the reduced data set and therefore lower compared to the full dataset (*Figure 2A–C*) with 0.89 g of ink. Overall, the data scattering was lower for the reduced data set.

Secondly, peroral dosing of PABA showed complete recovery in urine within 24 hours (94.75 ± 3.54%, cf. section *PABA metabolism after oral and intradermal administration*). Since PABA is fully dissolved in the tattoo ink, it can be assumed that it is freely available in the body, as was the case in peroral administration. Therefore, the sum of excreted PABA in urine corresponds to the systemically available fraction of the applied ink after tattooing. Hence, the applied ink per session (sys) was calculated from PABA recovered within 24 hours after the start of tattooing (*Table 2*). Similarly, applied ink was approximated by iodine excretion with subtraction of the iodine background from the 24 hours before tattooing (*Figure S1D–G*). For both iodine and PABA, the estimation of ink per session (sys) was about 25% lower compared to quantification via consumables (hypo).

**Table 2.** Comparison of mean ink exposure calculated from study data and its 75^th^ percentile with data used by the European Chemical Agency (ECHA).

| Basis of calculation | Sample size | Ink per session [g] |  | Ink per area [mg<sub>ink</sub>/cm<sup>2</sup>] |  |
| --- | --- | --- | --- | --- | --- |
|  |  | Mean | 75<sup>th</sup> p. | Mean | 75<sup>th</sup> p. |
| Ink per session (hypo) | n = 22 | 0.89 | 1.25 | 12.45 | 17.44 |
|  | (n = 13, recovery >25%) | 0.78 | 1.06 | 10.56 | 13.82 |
| Ink per session (sys, PABA) | n = 24 | 0.19 | 0.29 | 2.62 | 3.78 |
| Ink per session (sys, iodine) | n = 24 | 0.25 | 0.39 | 3.37 | 3.67 |
| Mean ink per session (sys, PABA/iodine) | n = 48 | 0.22 | 0.31 | 2.99 | 3.67 |
| ECHA calculation | n = 9 |  | 4.31 |  | 14.36 |

Note: 75<sup>th</sup> p.= 75<sup>th</sup> percentile, PABA= 4-aminobenzoic acid, sys= systemic available ink, hypo= hypothetically applied ink.

The ink per area (sys, PABA) is more positively correlated with red tattoos than black tattoos (*Figure 2D*). Likewise, a negative correlation with tattooed area was demonstrated for both colours (*Figure 2E*). An additional multiple factor analysis showed a minor connection between ink per area (sys, PABA), urine excretion and body fat (*Supplemental Material 6*).

The ECHA used the 75^th^ percentile of previously available data to calculate the ink per area, which resulted in an estimation of 14.36 mg_ink_/cm^2^. Thus, we also applied the 75^th^ percentile to our data set (*Table 2*). On the basis of the ink per session (hypo), our 13.82 mg_ink_/cm^2^ for the reduced data set with a recovery > 25% were close to the ECHA estimations. However, when ink per session (sys) was applied, the mean 75^th^ percentile was 3.67 mg_ink_/cm^2^ (*Table 2*).

### Metabolite profile and plasma kinetics

Several PABA metabolites were identified in the subjects’ urine. ACD was quantified in all subjects, whereas 4-aminohippuric acid (PAHA) was only above the limit of detection (LOD) in 15 subjects (*Supplemental Excel File*). The quantifiable metabolites of PABA were predominantly in the form of ACD (41.76 ± 11.15%), with only a small percentage in the form of PAHA (0.79 ± 0.76%)(*Figure S2*). Additionally, metabolites that could not be quantified directly, such as 4-acetamidobenzoic acid glucuronide (ACD-GlcA) and 4-acetamidohippuric acid (ACHA), were detected based on their monoisotopic masses (355.09 g/mol for ACD-GlcA and 236.08 g/mol for ACHA) and the corresponding extracted ion chromatograms (*Figure S3*).

In plasma, iodine and PABA metabolite ACD were detected, however ACD concentrations were close or below the limit of quantification (LOQ) or LOD (*Supplemental Excel File, Figure S4)*. This was particularly the case for subjects with low total PABA excretion in urine.

### PABA metabolism after oral and intradermal administration

Peroral administration resulted in an average PABA urine recovery of 94.75 ± 3.54% (*Table S1*), which is considered as complete excretion. For comparison between peroral and intradermal administration, data of three tattoo study subjects were selected based on their similar applied ink per session (hypo), resulting in a similar applied amount of PABA (48.21 mg, 36.00 mg, 38.82 mg). Also, their total PABA recovery in urine of 22.63 ± 1.84% was similar (*Table S1*).

As some metabolites could not be quantified, a peak area comparison was carried out. Peak areas are not necessarily related to concentrations since ionisation efficiencies of each metabolite may vary significantly. However, it allows for a general comparison of the metabolite profile of the two routes of administration. Therefore, all corresponding peaks were integrated and the ratio of the respective peaks to the total peak area of all metabolites in urine C–F was determined (*Figure 4*). After 24 hours, almost one fifth (18.83 ± 3.26%) of the peak areas correspond to ACHA when administered perorally, whereas intradermal application resulted in 3.20 ± 0.69%. The ACD content was higher after tattooing compared to peroral administration, which was already determined by quantification. No differences were observed for the ACD-GlcA areas.

**Figure 4.**
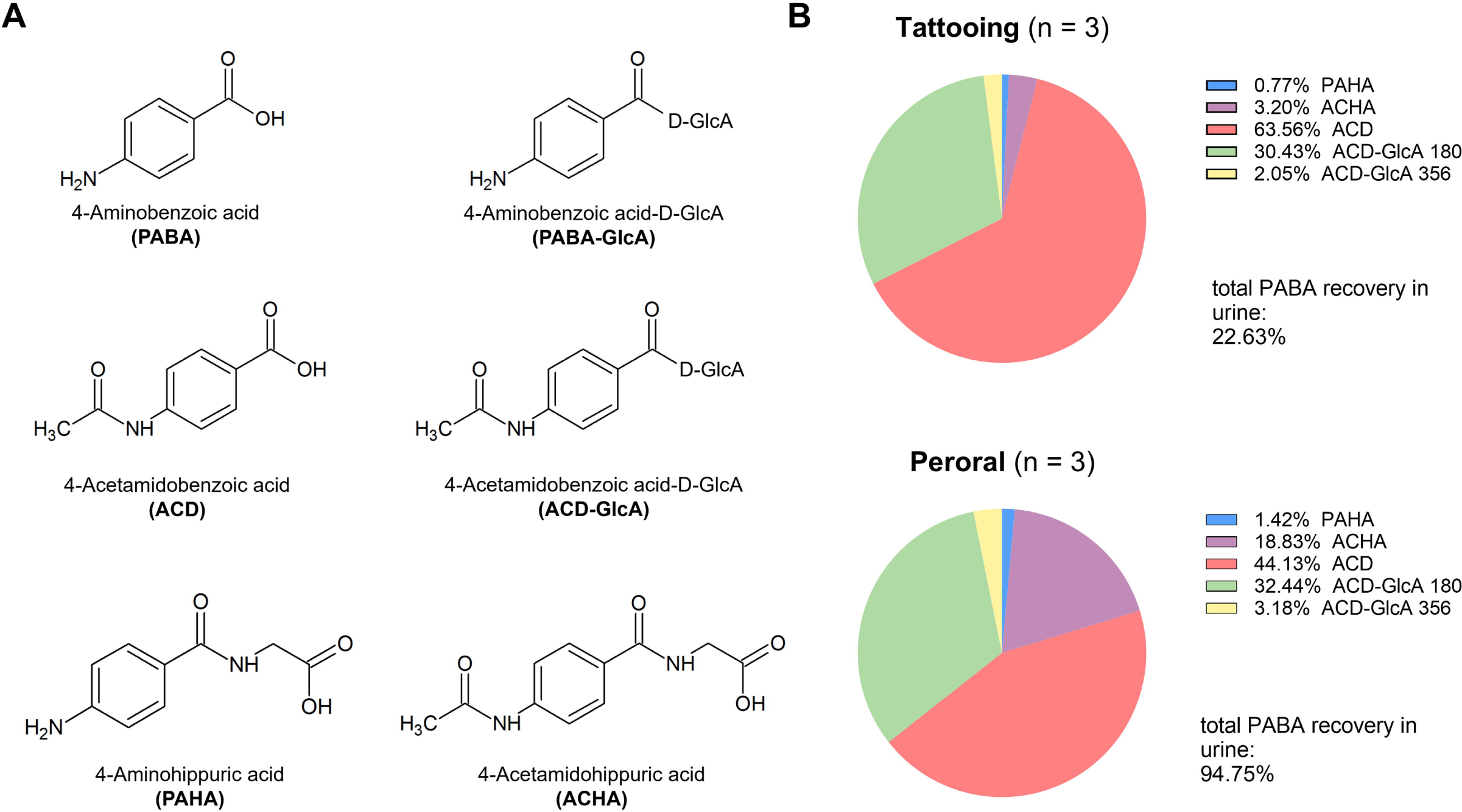
Analysis of 4-aminobenzoic acid and its metabolites. (A) Structural formulas of known PABA metabolites. (B) Comparison of the metabolite distributions in total urine (urine C–F) calculated from raw peak area ratios. Data are displayed for three tattoo study subjects (Black 8, Black 10, Red 5) and the three peroral PABA subjects for the first 24 h after tattooing.

### PABA metabolism in fibroblasts and monocyte-derived macrophages

Since our study results and the peroral comparison indicate a cutaneous first-pass effect during tattooing, the metabolic capacity to form ACD in tattoo-relevant skin cells was investigated. After 24 hours of incubation with PABA, both fibroblasts and monocyte-derived macrophages catalysed formation of ACD (*Table S2*). The samples were also screened for other known PABA metabolites via the corresponding extracted ion chromatograms in the obtained data sets, but no other metabolites were observed.

## Discussion

In this study, we obtained *in vivo* human exposure data on tattooing that are the first of their kind and represent a reasonable worst-case scenario. These data can be used to adapt risk assessments related to carcinogens and other toxic substances in tattoo and permanent make-up inks, thereby allowing for a better prediction of tattoo-associated health risks in the general population. Additionally, our data provide evidence for the capacity of skin cells to metabolise certain substances to an extent that affects parent compound-inherent toxic properties during tattooing, especially in the case of PAAs.

Iodine quantification in plasma was most successful since it is least affected by matrix effects and has a significantly lower LOD.^23^ Neither PEtOH nor its metabolite PAc were detected in plasma. Given that the concentration of the PABA metabolite ACD was already close to the LOD, it is reasonable that neither PEtOH nor PAc could be detected, as the PEtOH concentration in the ink was six times lower when compared to PABA and detection limits of PEtOH and PAc were slightly higher.^23^ Blood samples only reflect a specific time point, bearing the risk to miss the peak concentrations or any concentration above the LOD within the plasma kinetics. In urine samples, both PABA and PEtOH parent compounds were absent since they undergo fast metabolic conversion. Also, PABA-GlcA was not found despite previously reported as PABA metabolite.^38^ In case of PEtOH, *in silico*^39^ and animal experiments^40,41^ suggest a variety of possible metabolites, including those formed via oxidation, hydroxylation, sulfation or glucuronidation. In one human study, PEtOH was excreted in the form of both PAc (85%) and PAc conjugates (15%, n = 1), whereas another study on four subjects only detected PAc.^40,41^ A recent study by Eckert et al. identified 4-hydroxyphenoxyacetic acid as an additional metabolite in a lower ratio compared to PAc.^42^ In our study, no metabolite other than PAc was found. However, the PEtOH concentration used was much lower compared to the study by Eckert et al., hence it may be possible that 4-hydroxyphenoxyacetic acid was formed but below identification limits.

In our study, red tattoos correlated with higher use of ink but smaller tattooed areas when compared to black tattoos. This correlation can be explained by the fact that red tattoo designs mostly consisted of filled areas rather than shading and outlines which were more prominent in black tattoos. In the picture analysis, shaded areas were not considered. Therefore, tattoo design rather than ink colour plays an important role in the degree of ink exposure. Minor correlations of ink per area and total excreted urine volume were observed. With the small number of study subjects, the power of such analysis is limited. However, similar findings regarding increased excretion of iodine and other biomarkers with high urine volume have been reported previously.^43,44^

A subject’s wound dressing showed distinguishable red and black tattoo ink remains after 24 hours, indicating ink exudation through the damaged skin barrier. It is therefore most likely that not all of the ink applied will be absorbed by the body. Also, the recovery of tracers was extremely variable and ranged between 3.16 and 79.50%. Since all three tracers resulted in similar recoveries within each subject, analytical errors are unlikely to cause the low tracer recovery. An important source of errors is the indirect calculation of ink in consumables. Failing to collect all materials could lead to an underestimation of ink in consumables, leading to an overestimation of applied ink. Due to the variety of materials used by the artists that were not all tested during method validation, quantitation errors of the amount of ink adhered to consumables are certainly possible. Incomplete urine collection could also lead to an underestimation of excreted tracers. Factors influencing the absorption of ink into the body may include skin histology of different body parts, the tattoo artist’s technique or the wound healing process. Compound retention within the skin or the lymphatic system leading to a depot effect does not seem plausible in our view. Since the recovery rates in urine were similar for all three tracers, a possible depot effect in tissue could be only explained by physico-chemical differences, but is less likely to occur with small molecules (or ions) completely dissolved in the application media.^45^ Therefore, the most plausible factor responsible for the discrepancies in the systemic exposure levels might be transepidermal loss by wound healing. Accordingly, only a fraction of the hypothetically applied ink actually entered the body and that the fraction of soluble ink components absorbed into the body is likely overestimated using the hypothetically applied ink for exposure calculations.

This conclusion is also substantiated by our peroral *in vivo* data. PABA was completely excreted within 24 hours in the peroral setup, whereas PABA recovery during tattooing was only about 25% of the hypothetically applied ink. We concluded that the absorbed fraction of PABA and its metabolites would also be fully excreted during tattooing and calculated the systemically absorbed ink fraction from total PABA (and iodine) recovered in urine. Iodine was included because recovery results were similar to those of PABA. In this case, inaccuracies in consumable collection and iodine quantification are not relevant. Also, PABA is mainly excreted within the first hours after tattooing where study participants were still at the study centre, and where compliance in terms of complete urine collection is more reliable than at home. Therefore, only little effects on the exposure estimation are expected.

In the ECHA Annex XV restriction report, the 75^th^ percentile of the available data set was used to calculate a reasonable worst-case scenario, as these data reflected a high exposure situation only.^17^ In this study, only medium to large tattoos were included. Hence, according to our data, the 75^th^ percentile of 0.31 g ink per session (sys) displays a reasonable worst-case exposure level per tattoo session. This value is about 14-times lower than the 4.31 g per session previously estimated by the ECHA. Applying the exposure levels calculated from our data would have a significant impact on the risk assessment and health risk predictions of hazardous substances. For example, a market survey from 2022 analysed tattoo inks for their preservative concentrations and the highest PEtOH concentration found was 6475 µg/g_ink_ (about 0.65% PEtOH).^46^ Assuming that our calculated exposure value of 0.31 g ink per session is accurate, approximately 2 mg of PEtOH are introduced into the body during one tattooing session. However, according to REACH, only 0.01% PEtOH are allowed in tattoo inks due to its eye irritation properties. In cosmetics, 1% of PEtOH can be used and 0.5% is a common concentration to preserve injected pharmaceuticals.^25^ One of the highest findings of PAAs was 1775 µg/g_ink_ *o*-anisidine pre-REACH.^47,48^ However, PAA limits set by REACH are concentrations up to 5 µg/g_ink_. Here, a value of 0.31 g of ink would result in a maximum of 1.55 µg PAA entering the body when applying a REACH compliant tattoo ink preparation under reasonable worst-case conditions.^4^

The majority of research to date has been conducted in Europe and tattoo inks are currently more regulated in the European Union than in other regions. The United States are also seeking to increase consumer protection with the Modernization of Cosmetics Regulation Act of 2022 (MoCRA), which would also apply to tattoo inks.^49^ However, no quantitative limits for toxic substances in tattoo inks are defined in the proposed legislation.

In general, toxicological data on the intradermal application of substances are scarce. Therefore, transfer of data from other routes of administration (e.g., peroral) to the intradermal application route is justified to obtain an idea of the level of internal exposure. An important factor distinguishing tattooing from other absorption routes (e.g., peroral or intravenous) is cutaneous metabolism. Therefore, we compared the peroral intake of the tracer PABA to absorption via tattooing and investigated the metabolism of PABA in tattoo-relevant cells. The normalised peak area of the metabolite ACHA was more dominant in peroral administration when compared to intradermal administration, possibly due to the hepatic and gastrointestinal first-pass effect. No peak in the extracted ion chromatograms of PABA-GlcA was detected during chemical analysis, indicating that either this is not a favoured metabolic pathway at the concentrations administered in this study or that its signal was below background signal. When normalised to the total hydrolysed PABA in urine, the tattoo samples contained about twice as much ACD compared to peroral intake. Therefore, skin metabolism may play a role in detoxification of substances in inks that are known targets of the enzyme *N*-acetyltransferase 1. A detoxifying first-pass effect by *N*-acetylation of a genotoxic PAA (i.e., 2,4-toluenediamine) in skin and different skin cell types was previously described.^18^ Keratinocytes in the epidermis, which are known for their capacity to catalyse *N*-acetylation,^35,36,50,51^ may be in contact with the ink constituents during tattooing and in the early phase of wound healing. However, microscopic images of *post mortem* tattooed pig skin indicated that the majority of the ink is immediately deposited in the dermal layer with only minor direct contact to keratinocytes.^52^ Therefore, ACD formation during tattooing might be driven by fibroblasts and macrophages rather than keratinocytes. The comparison of peroral with intradermal and cell culture data indicates that the substances are likely to be metabolised in the skin during tattooing. A major difference between the two routes of administration is that tattooing delivers the dose unevenly over a longer period of time, depending on the motif, whereas peroral administration delivers the dose all at once. These differences certainly have an effect on the ratio of metabolites, as the routes of metabolism often depend on the dose of substance reaching the tissue. The quantitative impact on the detoxification of aromatic amines by this cutaneous first-pass effect compared to peroral administration is yet to be understood. Kinetic modelling might help to address this data gap in the future.

In conclusion, the present study fills the data gaps on systemic exposure to tattoo ink ingredients needed for health risk assessments. Given the loss of soluble and insoluble ink components via wound healing, future studies particularly should consider exudation. The data presented can be used to simulate the kinetics and internal exposure to these substances – ideally taking into account the cutaneous first-pass effect during tattooing as shown in this study. This may help to predict kinetics of substances that cannot be tested in human subjects due to their toxicological profile and to extrapolate toxicity data obtained from other sources (e.g., oral animal or *in vitro* data). Such additional data will contribute significantly to the efforts of our study to translate the kinetics and exposure data into health protection for consumers. The study data presented here should be used for discussion with competent authorities to develop harmonised exposure assessments and to evaluate tattoo-associated risks in the future.

## Data Availability

All data produced in the present study are available upon reasonable request to the authors

## Ethics approval

The study was approved by the ethics committee of the Charité – Universitätsmedizin Berlin (Berlin, Germany) under the proposal number EA4/085/21.

## Acknowledgements

We would like to thank Dr Annegret Blume, PD Dr Klaus Abraham and Dr Bernhard Monien for helpful discussions on the study design. We would also like to thank Dr Holger Hoffmann for the fruitful discussion on hydrolysis and our colleagues from Department 7 for providing cells and for the help with the realisation of peroral PABA metabolism. We would like to thank the entire study team of the Clinical Research Center for Hair and Skin Science for conducting the study, especially Dr Anna Lechner during study preparation. In addition, many thanks to the pharmacy of Charité-Universitätsmedizin Berlin, especially Dr Eric Woith and his team for spiking the tattoo inks with the tracers. Special thanks to the tattoo artists and study subjects, without whom this study would not have been possible.

## References

1. McGovern V. Metal Toxicity: Tattoos: Safe Symbols? Environmental Health Perspectives. 2005;113(9):A590. doi:10.1289/ehp.113-a590a

2. Kluger N, Seité S, Taieb C. The prevalence of tattooing and motivations in five major countries over the world. Journal of the European Academy of Dermatology and Venereology. 2019;33(12):e484–e486. doi:10.1111/jdv.15808

3. Borkenhagen A, Mirastschijski U, Petrowski K, Brähler E. Tattoos in der deutschen Bevölkerung – Prävalenzen, Soziodemografie und Gesundheitsorientierung. Bundesgesundheitsblatt - Gesundheitsforschung - Gesundheitsschutz. 2019/09/01 2019;62(9):1077-1082. doi:10.1007/s00103-019-02999-7

4. Commission Regulation (EU) 2020/2081 of 14 December 2020 amending Annex XVII to Regulation (EC) No 1907/2006 of the European Parliament and of the Council concerning the Registration, Evaluation, Authorisation and Restriction of Chemicals (REACH) as regards substances in tattoo inks or permanent make-up.

5. Agnello M, Fontana M. Survey on European Studies of the Chemical Characterisation of Tattoo Ink Products and the Measurement of Potentially Harmful Ingredients. In: Serup J, Kluger N, umler W, eds. Tattooed Skin and Health. S.Karger AG; 2015:0.

6. The Danish Environmental Protection Agency. Chemical substances in tattoo ink. Survey of chemical substances in consumer products no. 116. 2012.

7. Lehner K, Santarelli F, Vasold R, et al. Black Tattoos Entail Substantial Uptake of Genotoxicpolycyclic Aromatic Hydrocarbons (PAH) in Human Skin and Regional Lymph Nodes. PLOS ONE. 2014;9(3):e92787. doi:10.1371/journal.pone.0092787

8. Lehner K, Santarelli F, Vasold R, König B, Landthaler M, Bäumler W. Black tattoo inks are a source of problematic substances such as dibutyl phthalate. Contact Dermatitis. 2011;65(4):231–238. doi:10.1111/j.1600-0536.2011.01947.x

9. Regensburger J, Lehner K, Maisch T, et al. Tattoo inks contain polycyclic aromatic hydrocarbons that additionally generate deleterious singlet oxygen. Experimental Dermatology. 2010;19(8):e275–e281. doi:10.1111/j.1600-0625.2010.01068.x

10. Lim H-H, Shin H-S. Sensitive Determination of Volatile Organic Compounds and Aldehydes in Tattoo Inks. Journal of Chromatographic Science. 2017;55(2):109–116. doi:10.1093/chromsci/bmw163

11. Battistini B, Petrucci F, De Angelis I, Failla CM, Bocca B. Quantitative analysis of metals and metal-based nano- and submicron-particles in tattoo inks. Chemosphere. 2020/04/01/ 2020;245:125667. doi:10.1016/j.chemosphere.2019.125667

12. Forte G, Petrucci F, Cristaudo A, Bocca B. Market survey on toxic metals contained in tattoo inks. Science of The Total Environment. 2009/11/15/ 2009;407(23):5997-6002. doi:10.1016/j.scitotenv.2009.08.034

13. Foerster M, Ezzedine K, Ribet C, Zins M, Goldberg M, Schüz J. Tattoos: a new ancient carcinogen? ISEE Conference Abstracts. 2022;2022(1)doi:10.1289/isee.2022.P-0359

14. Schreiver I, Hesse B, Seim C, et al. Synchrotron-based ν-XRF mapping and μ-FTIR microscopy enable to look into the fate and effects of tattoo pigments in human skin. Scientific Reports. 2017/09/12 2017;7(1):11395. doi:10.1038/s41598-017-11721-z

15. Arbache S, Mattos EdC, Diniz MF, et al. How much medication is delivered in a novel drug delivery technique that uses a tattoo machine? International Journal of Dermatology. 2019;58(6):750–755. doi:10.1111/ijd.14408

16. Engel E, Santarelli F, Vasold R, et al. Modern tattoos cause high concentrations of hazardous pigments in skin. Contact Dermatitis. 2008;58(4):228–233. doi:10.1111/j.1600-0536.2007.01301.x

17. European Chemicals Agengy (ECHA). Annex XV Restriction Report. Substances in tattoo inks and permanent make up. 2017.

18. Grohmann L, Becker D, Rademann J, Ma N, Schäfer-Korting M, Weindl G. Biotransformation of 2,4-toluenediamine in human skin and reconstructed tissues. Archives of Toxicology. 2017/10/01 2017;91(10):3307-3316. doi:10.1007/s00204-017-1954-5

19. Bhaiya P, Roychowdhury S, Vyas PM, Doll MA, Hein DW, Svensson CK. Bioactivation, protein haptenation, and toxicity of sulfamethoxazole and dapsone in normal human dermal fibroblasts. Toxicology and Applied Pharmacology. 2006/09/01/ 2006;215(2):158-167. doi:10.1016/j.taap.2006.02.006

20. Wiegand C, Hewitt NJ, Merk HF, Reisinger K. Dermal Xenobiotic Metabolism: A Comparison between Native Human Skin, Four in vitro Skin Test Systems and a Liver System. Skin Pharmacology and Physiology. 2014;27(5):263–275. doi:10.1159/000358272

21. Strandt H, Voluzan O, Niedermair T, et al. Macrophages and Fibroblasts Differentially Contribute to Tattoo Stability. Dermatology. 2020;237(2):296–302. doi:10.1159/000506540

22. Baranska A, Shawket A, Jouve M, et al. Unveiling skin macrophage dynamics explains both tattoo persistence and strenuous removal. Journal of Experimental Medicine. 2018;215(4):1115–1133. doi:10.1084/jem.20171608

23. Kochs S, Schiewe S, Zang Y, et al. 4-Aminobenzoic acid, 2-phenoxyethanol and iodine used as tracers in a short-term in vivo-kinetics study for tattoo ink ingredients: Mass spectrometry method development and validation. Journal of Chromatography B. 2023/09/01/ 2023;1229:123891. doi:10.1016/j.jchromb.2023.123891

24. Bührer C, Bahr S, Siebert J, Wettstein R, Geffers C, Obladen M. Use of 2% 2-phenoxyethanol and 0.1% octenidine as antiseptic in premature newborn infants of 23–26 weeks gestation. Journal of Hospital Infection. 2002/08/01/ 2002;51(4):305-307. doi:10.1053/jhin.2002.1249

25. Meyer BK, Ni A, Hu B, Shi L. Antimicrobial preservative use in parenteral products: Past and present. Journal of Pharmaceutical Sciences. 2007/12/01/ 2007;96(12):3155-3167. doi:10.1002/jps.20976

26. Roy J, Carrier S. Acute Hepatitis Associated with Treatment of Peyronie’s Disease with Potassium Para-Aminobenzoate (Potaba). The Journal of Sexual Medicine. 2008/12/01/ 2008;5(12):2967-2969. doi:10.1111/j.1743-6109.2008.00918.x

27. Sharma RS, Joy RC, Boushey CJ, Ferruzzi MG, Leonov AP, McCrory MA. Effects of Para-Aminobenzoic Acid (PABA) Form and Administration Mode on PABA Recovery in 24-Hour Urine Collections. Journal of the Academy of Nutrition and Dietetics. 2014/03/01/ 2014;114(3):457-463. doi:10.1016/j.jand.2013.07.045

28. Nauman J, Wolff J. Iodide prophylaxis in Poland after the chernobyl reactor accident: Benefits and risks. The American Journal of Medicine. 1993/05/01/ 1993;94(5):524-532. doi:10.1016/0002-9343(93)90089-8

29. Le Guen B, Stricker L, Schlumberger M. Distributing KI pills to minimize thyroid radiation exposure in case of a nuclear accident in France. Nature Clinical Practice Endocrinology & Metabolism. 2007/09/01 2007;3(9):611-611. doi:10.1038/ncpendmet0593

30. World Health Organization (WHO). Use of potassium iodide for thyroid protection during nuclear or radiological emergencies. 2011.

31. Jackson AS, Pollock ML. Generalized equations for predicting body density of men. British Journal of Nutrition. 1978;40(3):497–504. doi:10.1079/BJN19780152

32. International Council for Harmonisation of Technical Requirements for Pharmaceuticals for Human Use. ICH guideline M10 on bioanalytical method validation and study sample analysis EMA/CHMP/ICH/172948/2019. https://www.ema.europa.eu/en/documents/scientific-guideline/ich-guideline-m10-bioanalytical-method-validation-step-5_en.pdf (assessed 13.03.2023).

33. European Medicines Agency. Guideline on bioanalytical method validation EMEA/ CHMP/EWP/192217/2009 Rev. 1 Corr. 2**. https://www.ema.europa.eu/en/documents/scientific-guideline/guideline-bioanalytical-method-validation_en.pdf (assessed 13.03.2023).

34. Foerster M, Dufour L, Bäumler W, et al. Development and Validation of the Epidemiological Tattoo Assessment Tool to Assess Ink Exposure and Related Factors in Tattooed Populations for Medical Research: Cross-sectional Validation Study. Original Paper. JMIR Form Res. 2023;7:e42158. doi:10.2196/42158

35. Bonifas J, Bloemeke B. N-acetylation of aromatic amines: Implication for skin and immune cells. Frontiers in Bioscience-Elite. 2015-01-01 2015;7(2):305-321. doi:10.2741/e733

36. Eilstein J, Léreaux G, Budimir N, Hussler G, Wilkinson S, Duché D. Comparison of xenobiotic metabolizing enzyme activities in ex vivo human skin and reconstructed human skin models from SkinEthic. Archives of Toxicology. 2014/09/01 2014;88(9):1681-1694. doi:10.1007/s00204-014-1218-6

37. Aparicio-Soto M, Riedel F, Leddermann M, et al. TCRs with segment TRAV9-2 or a CDR3 histidine are overrepresented among nickel-specific CD4+ T cells. Allergy. 2020;75(10):2574–2586. doi:10.1111/all.14322

38. Chan K, Miners JO, Birkett DJ. Direct and simultaneous high-performance liquid chromatographic assay for the determination of p-aminobenzoic acid and its conjugates in human urine. Journal of Chromatography B: Biomedical Sciences and Applications. 1988/01/01/ 1988;426:103-109. doi:10.1016/S0378-4347(00)81931-3

39. Hewitt NJ, Troutman J, Przibilla J, et al. Use of in vitro metabolism and biokinetics assays to refine predicted in vivo and in vitro internal exposure to the cosmetic ingredient, phenoxyethanol, for use in risk assessment. Regulatory Toxicology and Pharmacology. 2022/06/01/ 2022;131:105132. doi:10.1016/j.yrtph.2022.105132

40. SCCS Opinion on Phenoxyethanol. Scientific Comittee on Cosumer Safety. 2016. SCCS/1575/16.

41. Hartwig A, Commission M. 2-Phenoxyethanol [MAK Value Documentation, 2017]. The MAK-Collection for Occupational Health and Safety. 73-116.

42. Eckert E, Jäger T, Hiller J, Leibold E, Bader M, Göen T. Biotransformation and toxicokinetics of 2-phenoxyethanol after oral exposure in humans: a volunteer study. Archives of Toxicology. 2024/06/01 2024;98(6):1771-1780. doi:10.1007/s00204-024-03717-2

43. Johner SA, Shi L, Remer T. Higher Urine Volume Results in Additional Renal Iodine Loss. Thyroid®. 2010/12/01 2010;20(12):1391-1397. doi:10.1089/thy.2010.0161

44. Pinto J, Debowska M, Gomez R, Waniewski J, Lindholm B. Urine volume as an estimator of residual renal clearance and urinary removal of solutes in patients undergoing peritoneal dialysis. Scientific Reports. 2022/11/05 2022;12(1):18755. doi:10.1038/s41598-022-23093-0

45. Benedetti MS, Whomsley R, Poggesi I, et al. Drug metabolism and pharmacokinetics. Drug Metabolism Reviews. 2009/08/01 2009;41(3):344-390. doi:10.1080/10837450902891295

46. Famele M, Lavalle R, Leoni C, et al. Quantification of preservatives in tattoo and permanent make-up inks in the frame of the new requirements under the REACH Regulation. Contact Dermatitis. 2022;87(3):233–240. doi:10.1111/cod.14105

47. Jacobsen E, Tønning K, Pedersen E, Bernt, N, Serup J, Høgsberg T, Nielsen E. Chemical Substances in Tattoo Ink: Survey of Chemical Substances in Consumer Products. 2012. https://www2.mst.dk/udgiv/publications/2012/03/978-87-92779-87-8.pdf (assessed on 17.05.2024).

48. Fels P, Lachenmeier DW, Hindelang P, Walch SG, Gutsche B. Occurrence and Regulatory Evaluation of Contaminants in Tattoo Inks. Cosmetics. 2023;10(5):141. doi:10.3390/cosmetics10050141

49. U.S. Food and Drug Administration (FDA). Modernization of Cosmetics Regulation Act of 2022 (MoCRA).

50. Hu T, Khambatta ZS, Hayden PJ, et al. Xenobiotic metabolism gene expression in the EpiDerm™ in vitro 3D human epidermis model compared to human skin. Toxicology in Vitro. 2010/08/01/ 2010;24(5):1450-1463. doi:10.1016/j.tiv.2010.03.013

51. Oesch F, Fabian E, Oesch-Bartlomowicz B, Werner C, Landsiedel R. Drug-Metabolizing Enzymes in the Skin of Man, Rat, and Pig. Drug Metabolism Reviews. 2007/01/01 2007;39(4):659-698. doi:10.1080/03602530701690366

52. Hering H, Sung AY, Röder N, et al. Laser Irradiation of Organic Tattoo Pigments Releases Carcinogens with 3,3′-Dichlorobenzidine Inducing DNA Strand Breaks in Human Skin Cells. Journal of Investigative Dermatology. 2018/12/01/ 2018;138(12):2687-2690. doi:10.1016/j.jid.2018.05.031

