## Supplemental Figures and Tables for "Tat_BioV: Tattoo ink exposure and biokinetics of selected tracers in a short-term clinical study of 24 subjects"

*Archives of Toxicology*

### 1. Adjustments for absorbed fraction of tracers

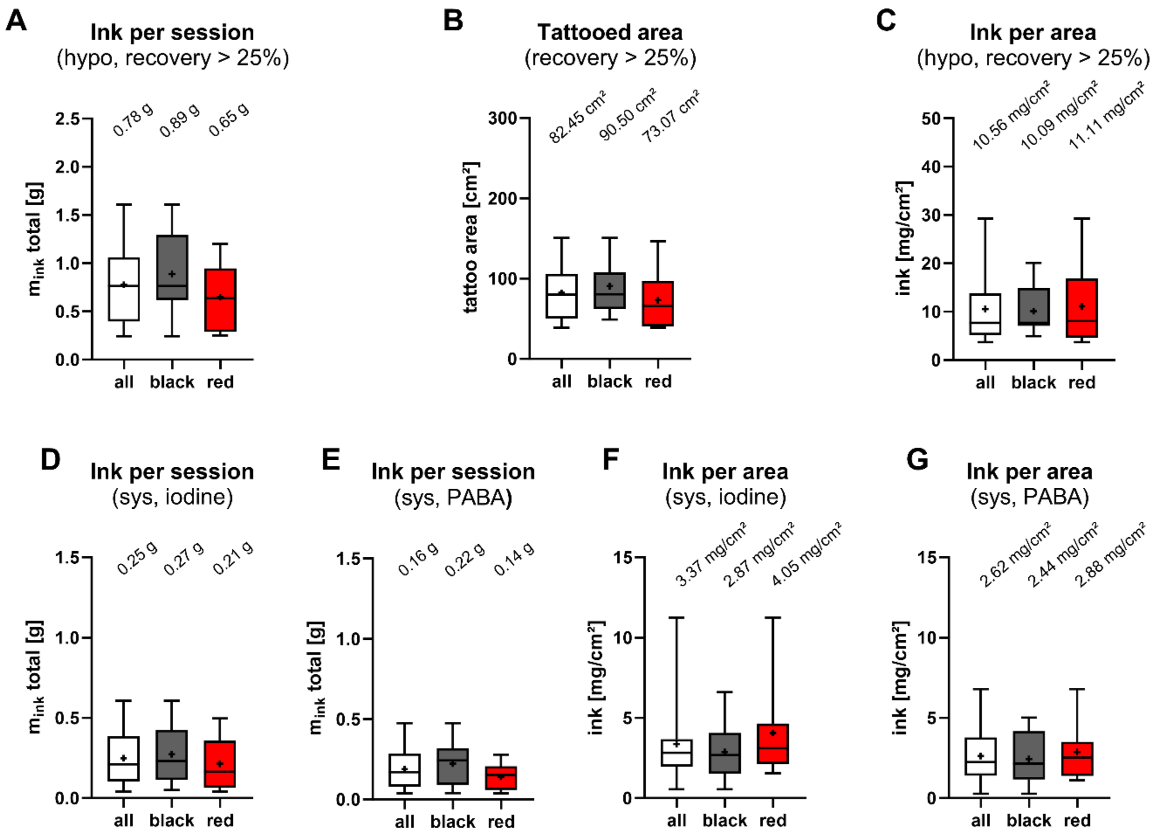


*Fig. S1: (A-C) Comparison of ink per session calculated from consumables, tattooed skin area and ink per surface of the reduced data set with mean urine tracer recovery above 25% (n = 13). D-G) Ink per session and corresponding ink per cm² calculated from the sum of recovered tracer substances (iodine or hydrolysed 4-aminobenzoic acid (PABA)) in urine (n = 24). The box and whisker plots show median (line) and mean (+) for all, black or red tattooed subjects.*

Regarding the tattooed area, the mean area decreases for black tattoos when the data set is reduced, while it increases for red tattoos. The average ink per area was lower for the total data set (all) and red tattoos, while it remained the same for black tattoos.

#### 2. Metabolic profile and plasma kinetics of selected subjects

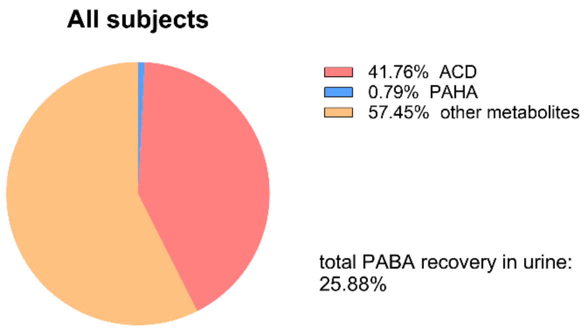


*Fig. S2. Metabolites of PABA quantified in total urine (C-F). ACD: 4-acetamidobenzoic acid; PAHA: 4-aminohippuric acid.*

The extracted ion chromatograms (EICs) of subject Black 8 displayed all PABA metabolites except PABA-GlcA (*Fig. S3*) in urine 4 h after starting the tattooing. The PAHA concentration in this subject was higher than in other subjects. Notably, ACD-GlcA also generated a signal in the EIC of ACD, in addition to its own $[M+H]^+$. The cleavage of the bond between glucuronide and ACD may be due to in-source decay during ionisation.

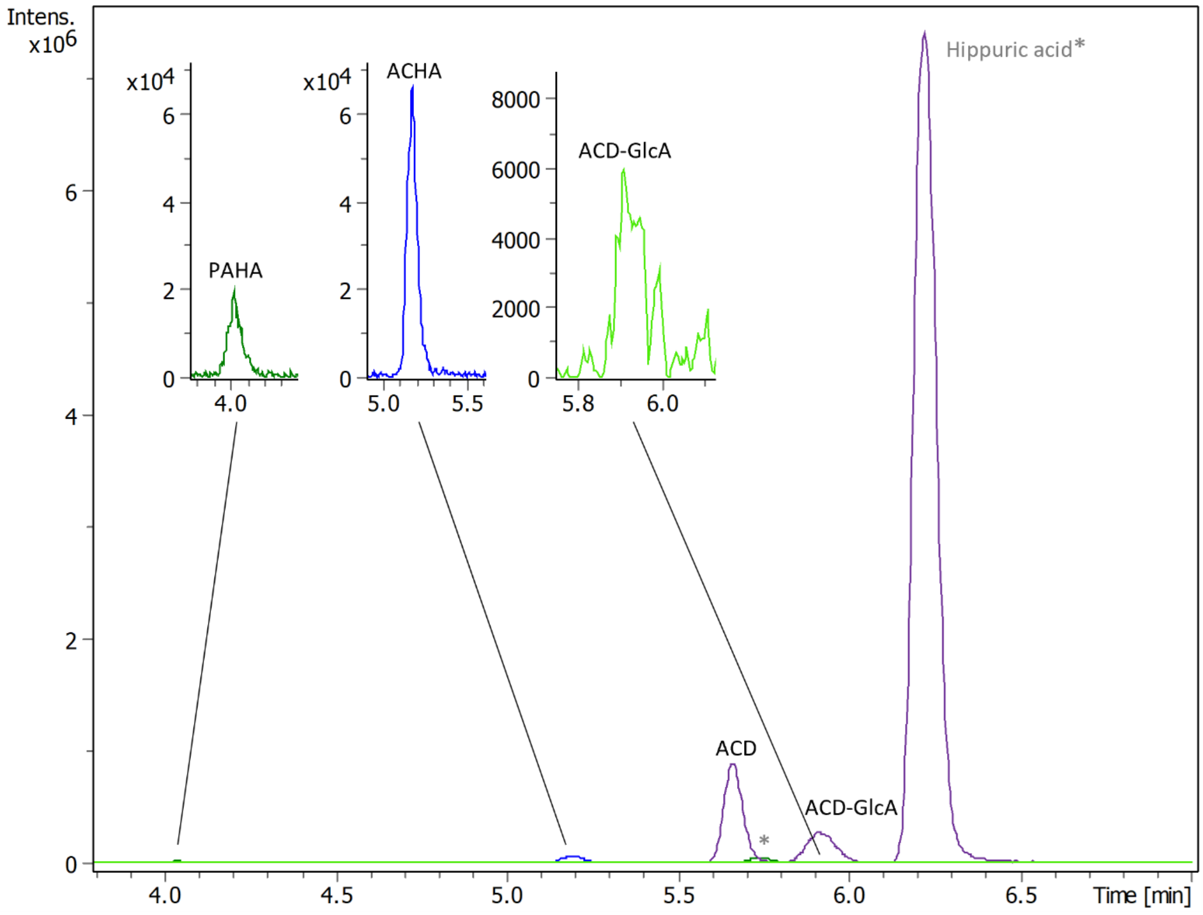


*Fig. S3. Analysis of 4-aminobenzoic acid (PABA) and its metabolites. Extracted ion chromatograms (EIC) of PABA metabolites in urine sample C of subject Black 8 with corresponding monoisotopic masses + H⁺. PAHA: EIC 195.076 ± 0.005; ACHA: EIC 237.087 ± 0.005; ACD and ACD-GlcA: EIC 180.066 ± 0.005; ACD-GlcA: EIC 356.098 ± 0.005. *Other background urine metabolites.*

The calibration curve for quantification was used to extrapolate all ACD concentrations between the LOQ and the LOD (*Supplementary Table 1*). Plasma kinetics of iodine and ACD of subject Black 5 are displayed for a session of 7 h 20 min (*Fig. S4*). A break was taken at around 4 h, which was reflected in the decrease in plasma concentrations of both substances. At 24 h, ACD was below the LOD and iodine declined to physiologically background levels, indicating elimination of the substances from blood. Neither PEtOH nor its metabolite PAc were found in plasma.

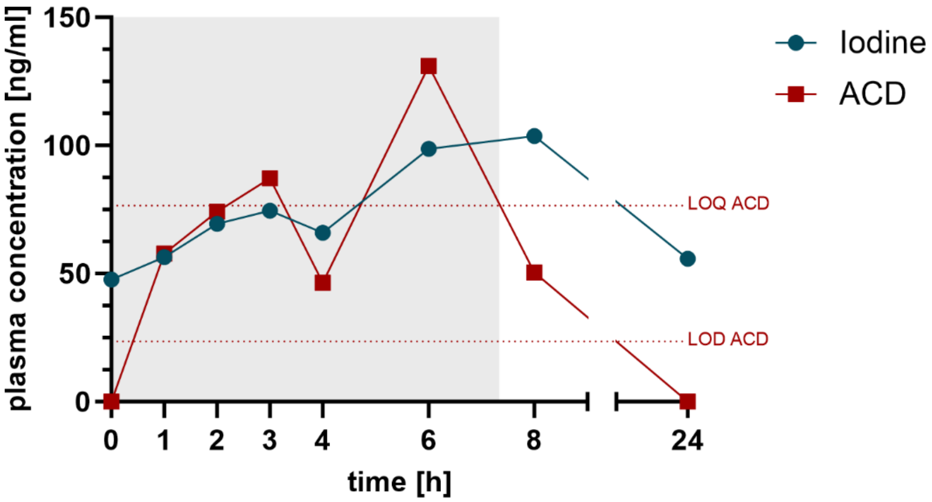


*Fig. S4. Plasma kinetics of iodine and 4-aminobenzoic acid metabolite 4-acetamidobenzoic acid (ACD) of subject Black 5 during the first 24 h after the start of tattooing. Limit of detection (LOD) and quantification (LOQ) for ACD are indicated in the graph, LOD and LOQ for iodine were 0.15 and 0.55 ng/ml, respectively. ACD concentrations between LOD and LOQ are extrapolated. The duration of the tattoo session was 7 h 20 min (indicated with grey background) with a break around 4 h.*

#### 3. PABA metabolism after oral and intradermal administration

For comparison between peroral and intradermal administration, data of three tattoo study subjects were selected based on their similar applied amount of ink per session (hypo), resulting in a similar applied amount of PABA (48.21 mg, 36.00 mg, 38.82 mg). The metabolites ACD and PAHA were quantified in urine C-F and the respective percentages from total PABA after hydrolysis was determined (*Table S3*). ACD formation was higher in tattooing compared to peroral administration.

*Table S1. Comparison of total urine recovery of 4-aminobenzoic acid (PABA) and quantified metabolites 4-acetamidobenzoic acid (ACD) and 4-aminohippuric acid (PAHA) excreted within 24 h after tattooing or peroral administration.*

| | | **Total PABA recovery [%]** | **Percentage ACD [%]** | **Percentage PAHA [%]** | **Percentage other metabolites [%]*** |
|---|---|---|---|---|---|
| Peroral | PABA I | 98.60 | 21.39 | 1.49 | 77.12 |
| | PABA II | 90.05 | 14.49 | 2.94 | 82.60 |
| | PABA III | 95.61 | 18.67 | 1.22 | 80.11 |
| | **Mean**$_{\text{Peroral}}$ | 94.75 | 18.18 | 1.88 | 79.94 |
| | **SD**$_{\text{Peroral}}$ | 3.54 | 2.84 | 0.75 | 2.23 |
| Tattooing | Black 8 | 20.14 | 37.21 | 2.04 | 60.76 |
| | Black 10 | 23.22 | 45.12 | 1.74 | 53.14 |
| | Red 5 | 24.52 | 33.58 | 0.46 | 65.96 |
| | **Mean**$_{\text{Tattooing}}$ | 22.63 | 38.64 | 1.42 | 59.95 |
| | **SD**$_{\text{Tattooing}}$ | 1.84 | 4.82 | 0.68 | 5.27 |

Note: SD = standard deviation; *Molar ratio calculated from total PABA after sample hydrolysis

### 4. PABA metabolism in fibroblasts and monocyte-derived macrophages

Both, fibroblasts and macrophages catalysed the formation of PABA metabolite ACD (*Table S4*). The results indicate that at a concentration of 1 µg/ml PABA, the enzymes were already saturated, as an increase of concentration did not result in increased formation of ACD. Since the macrophages were differentiated from blood of different donors, the concentrations exhibit a higher degree of variability.

*Table S2. Concentration of 4-acetamidobenzoic acid (ACD) after 24 h incubation in pooled human dermal fibroblasts and macrophages (fibroblasts: biological duplicate; macrophages: three different donors, all data points displayed as mean of technical triplicates).*

| | | $c_{ACD}$ [ng/ml] | | |
|---|---|---|---|---|
| | **$c_{PABA}$** | **0.1 µg/ml** | **1 µg/ml** | **10 µg/ml** |
| Fibroblasts | F.1 | 54.40 | 100.94 | n/a |
| | F.2 | 65.46 | 148.00 | 129.26 |
| | **Mean$_{Fibroblasts}$** | 59.93 | 124.47 | 129.26 |
| | **SD$_{Fibroblasts}$** | 5.53 | 23.53 | 0.00 |
| Macrophages | M.1 | 74.22 | 345.69 | 412.48 |
| | M.2 | 120.86 | 1165.07 | 1115.00 |
| | M.3 | 92.47 | 721.71 | 739.20 |
| | **Mean$_{Macrophages}$** | 95.85 | 744.16 | 755.56 |
| | **SD$_{Macrophages}$** | 19.19 | 334.89 | 287.04 |

Note: F = Fibroblasts, M = Macrophages, SD = standard deviation.
