## Supplemental Material for "Tat_BioV: Tattoo ink exposure and biokinetics of selected tracers in a short-term clinical study of 24 subjects"

*Archives of Toxicology*

#### Supplemental Material 1: Spiking of tattoo inks with selected tracer substances

##### Selected tracers

The tracer potassium iodide was used to determine the amount of ink applied to the dermis, since it is easy to track, not volatile at the used pH value and its quantification is not sensitive to metabolism or other processes. In medicine, iodide can be used in high concentrations to block the incorporation of the released radioactive iodine isotype by the thyroid hormone biosynthesis in case of nuclear incidents by saturating the responsible transport enzymes (Le Guen et al. 2007; Nauman and Wolff 1993; World Health Organization 2011). As iodide is physiologically present in human blood, its background levels have to be considered in the measurements and by setting the tracer concentrations accordingly (Nasterlack et al. 2016).

The second tracer used was 4-aminobenzoic acid (*p*-aminobenzoic acid, PABA). The potassium salt of this compound is used as a dietary supplement (Sharma et al. 2014) and for the treatment of Peyronie's disease (Weidner et al. 2005). However, concentrations used in this study were below levels used in drug products or dietary supplements (Sharma et al. 2014; Weidner et al. 2005). Here, it was chosen for its similarity to aryl compounds with an amino moiety, a common structure of primary aromatic amines (PAAs). Furthermore, PABA was selected for its high urine recovery rate of up to > 90 % 24 h after incorporation, which makes it a reliable indicator of the completeness of 24 h-urine collection (Bingham and Cummings 1983; Chan et al. 1988; Jakobsen et al. 2003). The metabolism of PABA is well-documented with potential metabolites including 4-aminobenzoic acid glucuronide (PABA-GlcA), 4-acetamidobenzoic acid (ACD), 4-acetamidobenzoic acid glucuronide (ACD-GlcA), 4-aminohippuric acid (PAHA) and 4-acetamidohippuric acid (ACHA) (Chan et al. 1988).

The third tracer was 2-phenoxyethanol (PEtOH). PEtOH is a preservative commonly used in cosmetic products and in some parenteral drugs (Bührer et al. 2002; Dréno et al. 2019), and sometimes also in tattoo inks.

##### Spiking of tracers

Commercially available black or red tattoo ink was selected by the tattoo artist and spiked with the tracers in a pharmacy unit. All tracers used were of pharmaceutical quality, i.e., according to European Pharmacopoeia (Ph. Eur.), or in form of a drug product. Due to the better solubility of the potassium salt (KPABA), POTABA capsules (Glenwood GmbH Pharmazeutische Erzeugnisse, Munich, Germany) were used in the case of PABA. PEtOH and potassium iodide were from Euro OTC Pharma GmbH (Bönen, Germany). The tracer concentrations were 30 mg/$g_{ink}$ PABA (38.33 mg/$g_{ink}$ KPABA), 5 mg/$g_{ink}$ PEtOH and 4.44 mg/$g_{ink}$ iodide (5.81 mg/$g_{ink}$ KI). The concentrations of iodide and PABA were selected to be below levels used in drug products but above detection limits in blood and urine (Weidner et al. 2005). PEtOH was used at a concentration for preservation in accordance to regulation (EC) No 1223/2009 (European Parliament 2009). To avoid contamination of the ink, the spiking was carried out in a cytostatic workbench according to DIN 12980 and EU-GMP cleanroom classification grade A, which was located in a grade C cleanroom. The microbiological burden was tested beforehand according to the European Pharmacopoeia. Tests according to chapter 2.6.12 (Microbiological examination of non-sterile products: microbial enumeration tests) and 2.6.13 (Microbiological examination of non-sterile products: test for specified microorganisms) were used to determine specific microorganisms (Council of Europe 2020). Neither the REACH restriction nor the German law on tattoo inks include specificities on the microbial status of the inks – according to the German Food and Feed Code the inks have just to be save (Lebensmittel- und Futtermittelgesetzbuch (English: German Food and Feed Code) 2005). As European and German regulations lack clarity regarding the microbial status of tattoo inks, other sources were taken into consideration. Swiss authorities concluded to tolerate a colony forming unit (CFU) of 10 per ml of ink and 100 CFU for opened ink bottles (Baumgartner and Gautsch 2011). The latter CFU is also specified for cosmetics for use in children under 3 years of age or on the eye or mucous membrane (EN ISO 17516:2014 2014). Here, only one test showed a low CFU count above the 10 CFU/ml limit of detection, which was not confirmed in the additional follow-up test for specified microorganisms. It was therefore concluded that the inks, also after spiking, were save to use in terms of their microbial status. The spiked ink was stored and transported at 2-8 °C and used within 24 h.

#### References


Baumgartner A, Gautsch S (2011) Hygienic-microbiological quality of tattoo- and permanent make-up colours. Journal für Verbraucherschutz und Lebensmittelsicherheit 6(3):319-325 doi:https://doi.org/10.1007/s00003-010-0636-5

Bingham S, Cummings JH (1983) The Use of 4-Aminobenzoic Acid as a Marker to Validate the Completeness of 24 H Urine Collections in Man. Clinical Science 64(6):629-635 doi:https://doi.org/10.1042/cs0640629

Bührer C, Bahr S, Siebert J, Wettstein R, Geffers C, Obladen M (2002) Use of 2% 2-phenoxyethanol and 0.1% octenidine as antiseptic in premature newborn infants of 23–26 weeks gestation. Journal of Hospital Infection 51(4):305-307 doi:https://doi.org/10.1053/jhin.2002.1249

Chan K, Miners JO, Birkett DJ (1988) Direct and simultaneous high-performance liquid chromatographic assay for the determination of p-aminobenzoic acid and its conjugates in human urine. Journal of Chromatography B: Biomedical Sciences and Applications 426:103-109 doi:https://doi.org/10.1016/S0378-4347(00)81931-3

Council of Europe (2020) European Pharmacopoeia 10th Edition.

Dréno B, Zuberbier T, Gelmetti C, Gontijo G, Marinovich M (2019) Safety review of phenoxyethanol when used as a preservative in cosmetics. Journal of the European Academy of Dermatology and Venereology 33(S7):15-24 doi:https://doi.org/10.1111/jdv.15944

EN ISO 17516:2014 (2014) Cosmetics - Microbiology - Microbiological limits.

European Parliament (2009) Regulation(EC) No 1223/2009 of the European Parliament and of the Council of 30 November 2009 on cosmetic products.

Jakobsen J, Pedersen AN, Ovesen L (2003) Para-aminobenzoic acid (PABA) used as a marker for completeness of 24 hour urine: effects of age and dosage scheduling. European Journal of Clinical Nutrition 57(1):138-142 doi:https://doi.org/10.1038/sj.ejcn.1601505

Le Guen B, Stricker L, Schlumberger M (2007) Distributing KI pills to minimize thyroid radiation exposure in case of a nuclear accident in France. Nature Clinical Practice Endocrinology & Metabolism 3(9):611-611 doi:https://doi.org/10.1038/ncpendmet0593

Lebensmittel- und Futtermittelgesetzbuch (English: German Food and Feed Code) (2005).

Nasterlack M, Drexler H, Hartwig A, Commission M (2016) Iodine and inorganic iodides [BAT Value Documentation, 2016] The MAK-Collection for Occupational Health and Safety. p 1535-1545

Nauman J, Wolff J (1993) Iodide prophylaxis in Poland after the chernobyl reactor accident: Benefits and risks. The American Journal of Medicine 94(5):524-532 doi:https://doi.org/10.1016/0002-9343(93)90089-8

Sharma RS, Joy RC, Boushey CJ, Ferruzzi MG, Leonov AP, McCrory MA (2014) Effects of Para-Aminobenzoic Acid (PABA) Form and Administration Mode on PABA Recovery in 24-Hour Urine Collections. Journal of the Academy of Nutrition and Dietetics 114(3):457-463 doi:https://doi.org/10.1016/j.jand.2013.07.045

Weidner W, Hauck EW, Schnitker J (2005) Potassium Paraaminobenzoate (POTABA™) in the Treatment of Peyronie's Disease: A Prospective, Placebo-Controlled, Randomized Study. European Urology 47(4):530-536 doi:https://doi.org/10.1016/j.eururo.2004.12.022

World Health Organization (2011) Use of potassium iodide for thyroid protection during nuclear or radiological emergencies.

#### Supplemental Material 2: Subject protocols

### Information on food and general rules

for the study

*Bioavailability of tattoo ink ingredients – analysis of blood and urine*

**The following applies to all rules: if it was not possible to comply with the rules before, during or after study participation, notification of the investigator/study physician is required.**

Medication & nutritional supplements:

Do not use **antibiotics or the antifibrotic drug POTABA®** for the duration of the study. **PABA** (4-aminobenzoic acid) and **iodide as nutritional supplements** should also be avoided. You may take other **nutritional supplements/vitamins and drugs** (including herbal drugs) **after consultation** with the investigator (study doctor), but they should be avoided if possible.

Stimulants:

**Alcohol and other toxicants or stimulants** should not be consumed during the 24 h before and after tattooing. Smoking (incl. e-cigarettes, heat-not-burn products, etc.) should be avoided as much as possible 1-2 h before, during, and 1-2 h after tattooing.

Food & Drinks:

Please do not drink **caffeinated beverages such as coffee, black and green tea, soft drinks containing caffeine or energy drinks** in the morning before, during, and 2-3 h after tattooing. Especially green tea and products containing green tea should be avoided completely. Avoid burned or heavily smoked foods (grill/oven/pan) as well as spinach.
**IMPORTANT:** Make sure that your meals are not excessively salted. Consumption of **seaweed (incl. sushi), fish or seafood** and unusually high amounts **of iodised table salt** should also be avoided.

Household products & cosmetics:

Since 2-phenoxyethanol can also be used as preservative in cosmetics, creams, cleaning products, hygiene products and soaps, do **not** use **your own cosmetic products, wound disinfection (especially Octenisept) and soap during the 48 h of the study period.** Please use the previously tested products contained in the wash bag provided beforehand and restrain from using any other.

Working with paints and similar products:

Any work with paint should not be carried out during the 24 h before and after tattooing and staying in rooms where paint has been used extensively should be avoided. This includes **wall paints, varnishes, spray paints, finger paints, art supplies, adhesives, clay (for pottery), putty, and other construction materials, as well as clay painting and related products.**

Other activities:

As with any fresh tattoo, protect it from sunlight and other UV sources until healing and **avoid sun exposure for 24 h after the tattooing took place**. In general, **avoid anything burned**. This includes sitting by a campfire or barbecue and for instance cleaning a barbecue grate and touching coals.

#### Supplemental Material 3: Analytical method description

##### Iodine quantification

For iodine quantification, plasma and urine samples as well as ink samples and consumables were analysed after sample preparation as previously described (Kochs et al. 2023). In short, plasma and urine samples were centrifuged at 3400 rcf for 15 min to remove suspended solids. Afterwards, these samples were then diluted by 1:50 (v/v) with final concentrations of 1% tetramethylammonium hydroxide (TMAH) in ultrapure water and 5 ppb tellurium as internal standard. TMAH 25% solution in $H_2O$ was acquired from VWR International GmbH (Darmstadt, Germany, catalogue number: 3316-35-250ML), tellurium solution (1003 mg/l ± 3 mg/l in 10% $HNO_3$ w/w, <0.1% HF w/w) was from Sigma Aldrich (Taufkirchen, Germany). Consumables were placed in heat-sealed homogenising bags (WhirlPak, Nasco Sampling, Madison, WI, USA, catalogue number: CEC5.1) with 1 l ultrapure water for 2 x 30 min on every side to extract iodide under shaking (250 rpm) before addition of the internal standard. The ink was diluted 1:5000 (v/v) in total with 1% TMAH and tellurium as internal standard. ICP-MS samples were analysed on an iCap Q instrument (Thermo Fischer Scientific, Bremen, Germany) with an SC4-DX configuration autosampler (Elemental Service & Instruments, Mainz, Germany) and operated with Qtegra software version 2.10.3324.131 (Thermo Fischer Scientific, Bremen, Germany), which was also used for data analysis.

##### HPLC-QTOF method

An HPLC-QTOF method was used to quantify PABA, PEtOH and their metabolites in plasma, urine and ink as previously described (Kochs et al. 2023). Isotope-labelled internal standards were utilised for quantification against a calibration curve in the corresponding matrix (plasma or urine). In brief, sample preparation included addition of internal standards, protein precipitation by ice-cold acetonitrile (Carl Roth GmbH & Co, Karlsruhe, Germany, catalogue number: HN40.2), centrifugation and further dilution with ultrapure water.

In addition, alkaline hydrolysis was performed to determine the total PABA content in urine. For this, 100 µl urine were shaken with 100 µl 10 M sodium hydroxide solution (Merck KGaA, Darmstadt, Germany, catalogue number: 1.06469.1000, lot: B1668469923) in a 1.5 ml reaction tube at 95 °C and 1000 rpm for 2 h (ThermoMixer C, Eppendorf, Hamburg, Germany). After cooling, the hydrolysate was neutralised with 60 µl acetic acid (Merck KGaA, Darmstadt, Germany catalogue number: A11350, Lot: 207034) and the processing was continued with 100 µl of the sample as described.[1]

For the analysis of 2-phenoxyacetic acid (PAc) in urine samples a standard addition method was used. A solution of PAc (TCI Deutschland GmbH, Eschborn, Germany, catalogue number: P0107, Lot: VZPKG) in acetonitrile (450 µg/ml) was diluted with ultrapure water to a concentration of 22.5 µg/ml. This solution was then used to dilute the different calibration levels, resulting in final concentrations for analysis of 0, 100, 250, 400, 550, 700, 850, and 1000 ng/ml. In a 1.5 ml-reaction tube, 20 µl of urine sample were added to 75 µl of ultrapure water and 50 µl of internal standard mix. 80 µl of the equivalently diluted calibration solution per calibration level was added and centrifuged at 4 °C and 14 000 rcf for 15 min after adding another 375 µl of ice-cold acetonitrile. The resulting supernatant was diluted 1:3 (v/v) with ultrapure water and measured. To calculate the concentration in each urine sample, the intersection point with the x-axis was determined. An HPLC system type Dionex UltiMate 3000 series (Fisher Scientific GmbH, Schwerte, Germany) was operated with a Luna C18 column (5 µm, 100 Å, 150 × 3 mm) attached to a precolumn from Phenomenex (Phenomenex Ltd. Deutschland, Aschaffenburg, Germany) and coupled to a maXis 4G QTOF-MS (Bruker Daltonik GmbH, Bremen, Germany). The operating system was Hystar 5.1 (Bruker Daltonik GmbH, Bremen, Germany) and

Bruker Compass DataAnalysis 5.2 and Bruker Compass QuantAnalysis 5.2 (Bruker Daltonik GmbH, Bremen, Germany) were used for data analysis.

**Reference**

Kochs S, Schiewe S, Zang Y, et al. (2023) 4-Aminobenzoic acid, 2-phenoxyethanol and iodine used as tracers in a short-term in vivo-kinetics study for tattoo ink ingredients: Mass spectrometry method development and validation. Journal of Chromatography B 1229:123891 doi:https://doi.org/10.1016/j.jchromb.2023.123891

#### Supplemental Material 4: Screening of cosmetic products for 2-phenoxyethanol

Subjects were required to follow a specific protocol, which included the use of PEtOH-free cosmetic products. The products were selected according to their labels and additionally analysed. The products included a deodorant spray (S01), shampoo (S02), shower gel (S03), toothpaste (S04) and cream (S05). A hand cream containing PEtOH on the label was used as a positive control (S00).

The concentration of PEtOH used in the study was 0.5% and falls within the limit of 1% set by Annex V of Regulation (EC) No 1223/2009. To test the method, the chosen cosmetic products were spiked with a concentration of 0.5% PEtOH (except for the positive control). To do so, a stock solution of 20% PEtOH in acetonitrile (Carl Roth GmbH & Co. KG, Karlsruhe, Germany) was used to spike the cosmetic products ad 0.5 g to reach a concentration of 0.5% PEtOH. Upon spiking the samples, they were vortexed for 1 min. The spiked in samples were diluted with 25 ml acetonitrile and were first shaken by hand for 1 min and afterwards vortexed for 1 min. The samples were filtered through polypropylene syringe microfilters (0.45 µmm, Thermo Fischer Scientific, Bremen, Germany) and further diluted by 1:200 (v/v). Finally, the spiked samples were again diluted by 1:200 (v/v) with acetonitrile into a vial for measurement. The final concentration of PEtOH was 500 ng/ml. In parallel, the stock solution was used to dilute a sample with a concentration of 500 ng/ml PEtOH in ultrapure water for semi-quantification.

Unspiked cosmetics samples were also measured, accordingly. Cosmetic samples, including the positive control (S00), were weighed at 0.5 g in a 50 ml centrifugation tube and prepared as described above. Sample S05 was additionally ultrasonicated for 15 min at 60°C. The samples were filtered through polypropylene syringe microfilters (0.45 µm) and either directly measured (S01, S02, S04, S05) or diluted by 1:200 (v/v) due to high background peaks in the case of S03 and in the case of S00 due to the present PEtOH content. The samples were analysed using the above-mentioned HPLC-QTOF-MS method in positive mode.

The results of the spiking indicate that the method did not significantly affect the results, as no substantial loss of PEtOH was observed (*Table S1*).

*Supplemental Material Table 1. Concentration of cosmetic products spiked with 2-phenoxyethanol.*

| Sample | PEtOH amount in product [%] | Deviation from nominal value [%] |
|---|---|---|
| S01 deodorant spray | 0.43 | - 14.11 |
| S02 shampoo | 0.58 | + 15.06 |
| S03 shower gel | 0.57 | + 14.99 |
| S04 toothpaste | 0.49 | - 1.80 |
| S05 cream | 0.52 | + 3.16 |

All samples except for the positive control S00 and S04 were PEtOH-free. However, the concentration of S04 was very low, as it was semi-quantified as 0.000275% in the product, which can be considered neglectable. The concentration of the positive control S00 was 0.35%.

#### Supplemental Material 5: Cell culture

Pooled human dermal fibroblasts from CELLnTEC advanced cell systems AG (Bern, Switzerland, catalogue number: HDfp, Lot: MC1904099) were cultured in Dulbecco's Modified Eagle's Medium/F12 1:1 (DMEM/F12) with L-glutamine, 1.2 g/l $NaHCO_3$ and without phenol red from PAN-Biotech GmbH (Aidenbach, Germany, catalogue number: P04-41650). The medium was supplemented with 5% fetal calf serum (FCS) from Sigma Aldrich (Taufkirchen, Germany, catalogue number: S0615-500ML) and 1% Penicillin/Streptomycin (100 U/ml) from PAN-Biotech GmbH (Aidenbach, Germany, catalogue number: P06-07100). The cryopreserved cells were cultured for at least two weeks before further experiments were started. Cell incubation conditions were constant at 37 °C, 5% $CO_2$ and ≥90% humidity.

Monocyte-derived macrophages were isolated from buffy coats of three different donors as previously described (Aparicio-Soto et al. 2020). After isolation of the peripheral blood mononuclear cells (PBMCs), the PBMCs were seeded in a 12-well plate ($8.75 \times 10^5$ cells/well) in Roswell Park Memorial Institute (RPMI) 1640 medium without L-glutamine and with 2.0 g/l $NaHCO_3$ and phenol red from PAN-Biotech GmbH (Aidenbach, Germany, catalogue number: P04-17500). The medium was supplemented with 1% Penicillin/Streptomycin (100 U/ml), 10% FCS and 1% L-glutamine (200 mM, PAN-Biotech GmbH, Aidenbach, Germany, catalogue number: P04-80100). After allowing the cells to adhere for 1 h, they were washed with Dulbecco's phosphate-buffered saline (DPBS) without $Mg^{2+}$ and $Ca^{2+}$ (PAN-Biotech GmbH, Aidenbach, Germany, catalogue number: P04-36500) to remove all non-adherent cells. Cells were cultured with human recombinant macrophage colony-stimulating factor (rHuM-CSF, final concentration 25 ng/ml) from Biomol GmbH (Hamburg, Germany, catalogue number: 60530, Lot: 1414) for one week. As RPMI medium contains PABA as supplement, the treatment with PABA was done in DMEM/F12 after washing with DPBS. Cell incubation conditions were constant at 37 °C, 5% $CO_2$ and ≥90% humidity.

##### Reference

Aparicio-Soto M, Riedel F, Leddermann M, et al. (2020) TCRs with segment TRAV9-2 or a CDR3 histidine are overrepresented among nickel-specific CD4+ T cells. Allergy 75(10):2574-2586 doi:https://doi.org/10.1111/all.14322

#### Supplemental Material 6: Multiple factor analysis (MFA) of study data

To identify further factors that could possibly influence the amount of ink per area, we performed an MFA using the package “FactormineR” in R (version 4.3.1). This type of analysis has two main advantages: (1) It is possible to include categorical parameters together with continuous parameters and (2) parameters can be grouped according to their inherent structure. Here, duration of the tattoo session and area were grouped to “tattoo size”; body height, weight and fat were grouped into “personal data” (cf. groups and other parameters in *Table S2*). Since the differentiation between a “low” and “high” amount of used ink per area was of high interest, we assigned a categorical parameter according to amount of ink per area. The limit of 3.5 $mg_{ink}/cm^2$ was set according to the shape of the violine plot (*Figure S2A*). The following data were calculated with the total ink per session (sys, PABA) recovered in urine C-F.

*Supplemental Material Table 2: Grouping strategy for multiple factor analysis (MFA) of given parameters. Tattoo colour and ink per area were used as categorial parameters with the categories black or red and low or high, respectively.*

| Parameter | Unit / Category | Parameter type | MFA Group |
|---|---|---|---|
| Tattoo colour | red / black | categorical | Tattoo colour |
| Session duration | min | continuous | Tattoo size |
| Tattooed area | cm² | continuous | |
| Body height | cm | continuous | Personal data |
| Body weight | kg | continuous | |
| Body fat | % | continuous | |
| Urine volume (C-F) | ml | continuous | Amount of urine |
| Ink per session | g | continuous | Ink per session |
| Ink per area | low / high | categorical | Ink per area |

The MFA separates samples according to independent dimensions, or principal components, and defines the extent to which a certain parameter contributes to this dimension. Plotting subject data according to dimension 1 and dimension 3, a clear separation of “low” and “high” amount of ink per area can be observed (*Figure S2B,C*). Interestingly, the same dimensions help to distinguish the individual samples according to the tattoo colour, supporting our initial finding. To better understand which parameters contribute to separation of individual samples, we analysed the contributions of single parameters to the dimensions (*Figure S2D*). The main factors influencing dimension 1 were total ink used per session and total volume of urine. For dimension 3, tattooed area and body fat contributed most. The other dimensions identified by MFA did not separate individual samples in relation to ink per area or the tattoo colour. A positive correlation of ink per area and total urine volume C-F may also be interpreted when looking at their correlation plot (*Figure S2E*), meaning that more PABA might be excreted with higher total urine volume. A negative correlation between the amount of ink per area and body fat might be contemplated. Correlations remain speculative due to the limited number of study subjects.

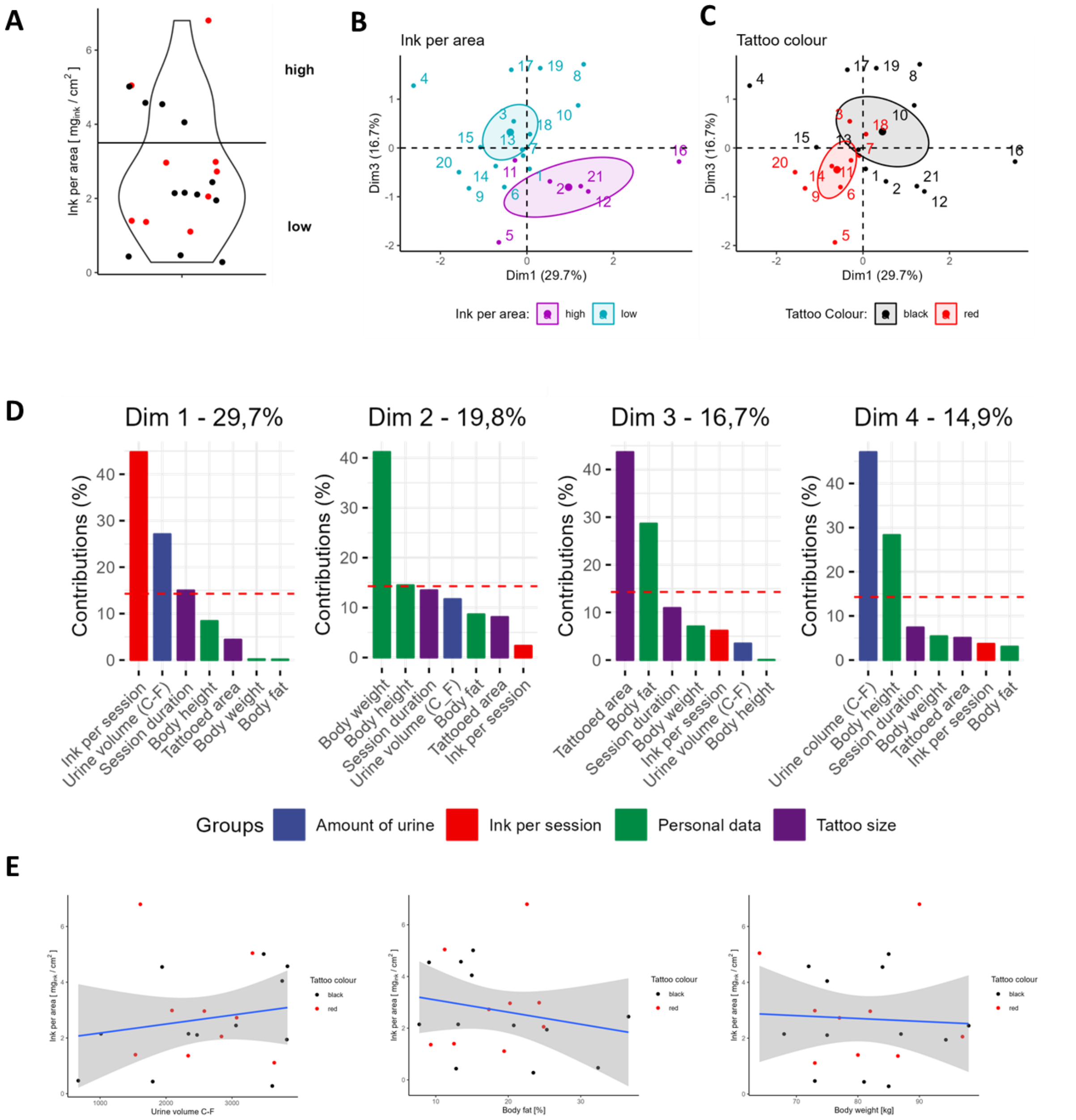


*Supplemental Material Figure 1. Correlation plots and multiple factor analysis (MFA) of study data. A: Violine Plot of ink per area. B,C: Separation of study data through MFA in the dimension 1 and 3 with indicated categorial parameters ink per area and tattoo colour. D: Single parameters determining the four dimensions. Percentage of influence on data separation of each dimension is indicated above the graphs. E: Correlation of the amount of ink per area (estimated from total 4-aminobenzoic acid excretion) against the ink per session, body fat and body weight, respectively.*
